# Spatio-temporal surveillance and early detection of SARS-CoV-2 variants of concern: a retrospective analysis

**DOI:** 10.1101/2023.07.06.23292295

**Authors:** Massimo Cavallaro, Louise Dyson, Michael J. Tildesley, Dan Todkill, Matt J. Keeling

## Abstract

The SARS-CoV-2 pandemic has been characterized by the repeated emergence of genetically distinct virus variants of increased transmissibility and immune evasion compared to pre-existing lineages. In many countries, their containment required the intervention of public health authorities and the imposition of control measures. While the primary role of testing is to identify infection, target treatment, and limit spread (through isolation and contact tracing), a secondary benefit is in terms of surveillance and the early detection of new variants. Here we study the spatial invasion and early spread of the Alpha, Delta, and Omicron (BA.1 and BA.2) variants in England from September 2020 to February 2022 using the random neighbourhood covering (RaNCover) method. This is a statistical technique for the detection of aberrations in spatial point processes, which we tailored here to community PCR (polymerase-chain-reaction) test data where the TaqPath kit provides a proxy measure of the switch between variants. Retrospectively, RaNCover detected the earliest signals associated with the four novel variants that led to large infection waves in England. With suitable data our method therefore has the potential to rapidly detect outbreaks of future SARS-CoV-2 variants, thus helping to inform targeted public health interventions.

## 1 Introduction

With more than 600 million confirmed cases and more than 6 million associated deaths worldwide by November 2022, the COVID-19 pandemic caused by SARS-CoV-2 has proven to be a substantial threat to global public health [1]. Its community-level burden is revealed by epidemiological surveillance, that is the systematic collection of health-related data and the real-time monitoring of trends of incidence through time [2]. Although testing is generally performed for patient-centred and public health reasons, the data on the number, location and type of infection is also of central importance as it provides information useful for the implementation of effective interventions to mitigate the overall burden on public health [3]. Mathematical and statistical modelling is often a critical part of this analysis, enabling us to translate the raw data into a projection of current and future trends with associated uncertainty bounds [4, 5]. While vaccination campaigns have decreased the overall risk of severe outcomes for COVID-19 cases and many countries have been phasing out restrictions since 2022, ongoing active surveillance is still required due the rapid emergence of SARS-CoV-2 mutants with new, potentially threatening, phenotypic properties [6]. SARS-CoV-2 surveillance includes identifying new variants, assessing their outbreak potential, and timing and targeting interventions to geographical regions who need them the most [7]. This last step is extremely important as interventions, either pharmaceutical or non-pharmaceutical, are most effective when enacted early and yet they are both economically and socially costly, and should therefore be avoided where and when unnecessary. The quest for tools to detect increases in the number of infections had led the research community to develop a number of statistical surveillance systems (as reviewed in [8–11]), and, more recently, model-independent methods rooted in the theory of dynamical systems [12–15].

In this paper, we retrospectively apply a spatio-temporal statistical surveillance method called RaNCover (random neighbourhood covering) to study the relative prevalence of SARS-CoV-2 variants and flag the locations where a new variant was set to overtake previous lineages. RaNCover was recently developed to monitor invasive Group-A Streptococcal disease (iGAS) cases [16], but as we demonstrate here, can be easily applied to other epidemiological situations to detect a change in the pattern of cases.

In general, the detection of a change in surveillance data is a two-step process: first a simple model is fitted to predict the expected behaviour and uncertainty (e.g., predicting the distribution of the number of cases) at each location and time; secondly these projections are then statistically compared to the observed counts. If the observation is above a specific percentile of the prediction interval – a situation which we call exceedance – then an alarm is raised [17]. One of the major challenges encountered in the development of an effective surveillance system is that available data are often patchy, with missing information in certain geographical locations and time intervals, and with varying sampling efforts. Dealing with insufficient data is particularly common at the beginning of an outbreak (of a new pathogen or new variant), when the number of cases is still low but is growing fast. Such sparse observations are often prohibitive when monitoring local trends and are subject to strong fluctuations that can potentially trigger many false positive alarms. One approach to this issue is to improve the prediction of the observed number of cases in a given space-time region by integrating the information available from nearby regions [18]. The random neighbourhood covering method introduced in [16] is an alternative approach: instead of improving model prediction in a spatio-temporal region, it aggregates information from multiple weak exceedance tests performed in neighbouring areas to improve the final warning signal relative to an area of interest. In other words, while the individual tests performed on single locations are likely to perform poorly, their consensus estimates have far better properties – an idea which is commonly exploited in ensemble statistical learning [19].

Here we considered SARS-CoV-2 polymerase-chain-reaction (PCR) test results collected in routine community testing in England (known as Pillar-2 testing) from August 2020 to February 2022. PCR results are obtained with a short delay (a few days in general) after testing and thus they offered a good opportunity to perform regular population-level surveillance. While these tests are principally used to inform about the aetiology of infection (distinguishing SARS-CoV-2 infection from other respiratory pathogens), the sum of all positive tests is a useful indicator of the scale of the epidemic. Here we focused on testing performed with the ThermoFisher TaqPath PCR Kit. This assay is built with redundancy as it targets three highly-specific regions of the SARS-CoV-2 genome, including a region in the S-gene (the gene that encodes the spike protein). When a sample fails to amplify one or two of the three targets, it can still be attributed to SARS-CoV-2, but the failure may also signal the presence of a SARS-CoV-2 variant: the Alpha and Omicron BA.1 variants fail to trigger the S-gene targeting element of the TaqPath system, whereas wildtype, Delta, and Omicron BA.2 do lead to S-gene positive tests [20, 21]. While whole genome sequencing is necessary to identify variants with certainty, S-gene detection has allowed a rapid assessment of the invasion of novel variants over time – without the delays (and expense) that are associated with genotyping (although some care is needed when the sample has a low viral concentration as indicated by a high cycle threshold, *Ct*, value). Historically, these data permitted the observation of SARS-CoV-2 Alpha variant spreading and becoming the dominant lineage in few months after being first detected in September 2020 [22, 23]. At the time, the data on S-gene detection alone was unlikely have offered the means to raise early alarms on the invading variant. However, with the benefit of hindsight, we can now search for early warning signals in these data and study how timely invasions can be detected in the presence of general data on SARS-CoV-2 variants. Later, the Alpha variant was out-competed by the Delta variant which became dominant in England by late May 2021 [24]. A third group of variants with both increased transmissibility and immune escape, called Omicron, were first detected in England at the end of September 2021 [25]. Each of these novel variants (Alpha, Delta, and Omicron) grew at a rate higher than those preceding, thus causing three major waves of infection during the study time.

Variants are defined by their genetic characteristics, and are initially labelled as variants of interest or variants of concern by public health agencies, such as the World Health Organization (WHO) and the UK Health Security Agency (UKHSA) in United Kingdom, based on their epidemiological characteristics. Variants of interest (VOIs) are those with genetic changes expected to increased COVID-19 transmissibility, severity, and diagnostic or therapeutic escape and actually observed to cause significant community transmission in multiple countries or other epidemiological repercussions, thus suggesting an emerging risk to global public health. Variants of concern (VOCs) are those where there are further detrimental changes in COVID-19 epidemiology, clinical disease presentation, or in the effectiveness of public health measures and diagnostics. As such variants are often first classified as VOIs before becoming VOCs, although not all will lead to a major increase in cases. The Alpha, Delta, and Omicron variants were designated as VOCs by the WHO and the UKHSA [7, 26–28]. However, detecting a new variant is only a part of the surveillance effort as policy-advisors need summarised information, such as how quickly and in which regions the variant is spreading. Here, we analysed the historical invasions of four variants – Alpha, Delta, Omicron BA.1, and Omicron BA.2 – in England with RaNCover, highlighting the Lower Tier Local Authority (LTLA) districts with exceedingly high levels of a variant as time progresses, and thus anticipating country-wide waves. The application of this approach in the future has the potential to provide a rapid assessment of a variant’s epidemiological potential and could inform early targeted interventions.

## 2 Methodology

Here we outline the two basic steps in the RaNCover methodology. We first describe the simple predictive model used to estimate the distribution of SARS-CoV-2 positive samples that are S-gene positive/negative each day in each Lower Tier Local Authority (LTLA). In brief, we estimate the number of S-gene positive/negative samples based on the number of tests in a given location on a given day and the recent historic proportions. Second, we focus on the comparison of this prediction with the observed data and the formulation of a warning score. We compare the true number of S-gene positive/negative in multiple overlapping regions and times around our point of interest, and compare each to our projections; warning scores are raised if the true values are significantly above our projections. A more detailed and technical description is given in the following two subsections.

### 2.1 Baseline levels of positive and negative tests

We considered SARS-CoV-2 positive tests in England performed with the ThermoFisher TaqPath system, an assay based on reverse-transcription polymerase chain reaction (RT-PCR). Detection of S-gene was used as a proxy to identify a variant, and S-gene positive and negative count data were aggregated by a patient’s LTLA as a proxy for location. In general LTLAs contain between 50,000 and 400,000 inhabitants, although the Isles of Scilly is the smallest LTLA with under 2,500 residents while Birmingham with 1.1 million residents is the largest. To assess the potential for a variant to be concerning, we measured the exceedance in the number of S-gene positive and S-gene negative cases with respect to a baseline level, which we derived as follows. For a given LTLA location *i* and day *t*, the aggregated data consists of the total number *N*_*i,t*_ of strongly positive community PCR tests using the TaqPath system (with a cycle threshold less than 30) and the number *M*_*i,t*_ of these tests which are positive for the S-gene. The proportion of S-gene positive tests, *M*_*i,t*_*/N*_*i,t*_ which is a proxy of the prevalence of a variant, is denoted as *P*_*i,t*_. The day *t* corresponds to the date on which a swab sample was collected; in general, the TaqPath results were then available 1-2 days later, which would add a slight delay to any detection mechanism. The objective of our methodology is to find any deviations from a null model for *M*_*i,t*_ in the observed data. The null model does not need to describe the data accurately, but only formalise how the data would behave if the dynamics of new infections were stable. We therefore estimated the average proportion 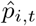 for our null model as a running average using values observed in the previous *T* = 15 days,

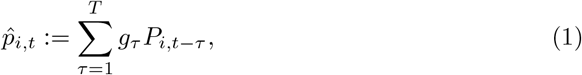

where the weight distribution *g*_*τ*_ is taken to be exponential with mean 5.5 days set by the approximate time-scale of infection duration [29–31]. This has the net effect of combining the observed statistics for each day and smoothing the test statistics over time. Conditioned on the total number of COVID-19 positive tests (*N*_*i,t*_) and the null model, the expected number of S-gene positive tests observed is 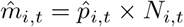. Analogous baseline values for the observed number of S-gene negative tests 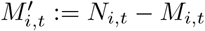 is 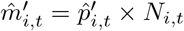, where 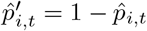.

### 2.2 Exceedance probability and random neighbourhood approach

We let

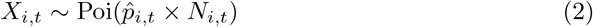

denote the Poisson random variable that describes the number of S-gene positive cases detected under the null model in location *i* at time *t*. Let us define a threshold value 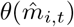 such that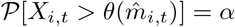 i.e., the probability that *X*_*i,t*_ exceeds the threshold is *α*. In other words, *θ* is the quantile function evaluated at 1 − *α*, see also electronic supplementary text S1. The basic exceedance criterion consists of raising a warning flag at location *i* at time *t* if the number of S-gene positive cases exceeds the threshold defined by the null model,

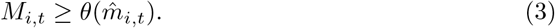

An analogous exceedance criterion for the number of S-gene negative cases is 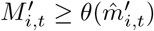. Exceedance probabilities under the Poisson model (2) and a more refined negative-binomial model are discussed in the electronic supplementary text S2.

A limitation of this naive approach is that there is a probability *α* of triggering a warning signal by chance even when the data come from the null model. A second limitation of the naive approach is that the criterion (3) is only applied to a single data point (one location and one time) whereas outbreaks are typically spatially and temporally extended; therefore it is desirable to test exceedance over a number of locations and days simultaneously, albeit it is impossible to know the outbreak range prospectively. We detect aberrations from the baseline by means of the random neighbourhood covering (RaNCover) approach of [16]. RaNCover tackles the limitations mentioned above by considering the exceedances over many overlapping spatio-temporal sets (chosen to be cylinders whose heights and circular bases corresponding to the time and geography components, respectively) that cover a region of interest and its neighbours. By virtue of the Poisson assumption (2), the number of observed S-gene positive or negative cases in a spatio-temporal set *C* is also a Poisson random variable, with intensity given by the sum of the intensities at all points (*i, t*) *∈ C*, i.e.,

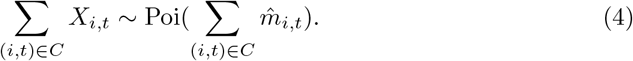

This allows us to flag a cylinder if

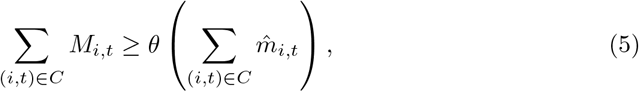

which is analogous to condition (3). A warning score *w*_*i,t*_ specific to the location (*i, t*) is then defined as the percentage of flagged cylinders over all sampled cylinders *C* such that (*i, t*) *∈ C*,

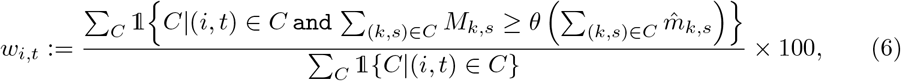

where 1 is the indicator function. Cylinder dimensions were randomly sampled, with uniformly drawn pairs of radii and heights (*ρ, h*) such that the cylinder volumes are constant (*π×h×ρ*^2^ = *c≈* 4066 day*×*km^2^). Upper limits to the heights and radii were heuristically set to 16 days and *≈* 36km, respectively, in order to cover short outbreaks over several LTLAs. Cylinder positions were also randomly chosen such that each cylinder contains an LTLA centroid location (this can be easily achieved by centering each cylinder at one of these locations and then shifting in space by an amount smaller than *ρ*; see also figure 1). To condition on cylinders not being empty, and therefore look at the neighbourhoods of identified test locations, the relation (2) is corrected to 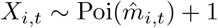 (see also [16]). The algorithm was recursively applied to each day within the study period. At each step, one of the bases of the cylinders was positioned on the plane corresponding to the current day, ensuring that the cylinders only extended towards the past and did not cover future time points. By doing so, all warning scores were computed using exclusively past information, thus emulating a real-time application. We drew 500,000 cylinders at each step. We expect that the statistical properties improve as the total number of cylinders and the sample size over which the sum in equation (7) is evaluated increases, but the estimation of the uncertainty associated with the warning scores requires additional theoretical studies (as an example, using confidence intervals for proportions, based on the number of cylinders covering a point event, as in the illustration S3 of reference [16], could underestimate uncertainties due to correlated observations in overlapping cylinders). On the other hand, as also discussed in [16], the algorithm appears to be robust to changes in cylinders’ numbers and dimensions.

**Figure 1.**
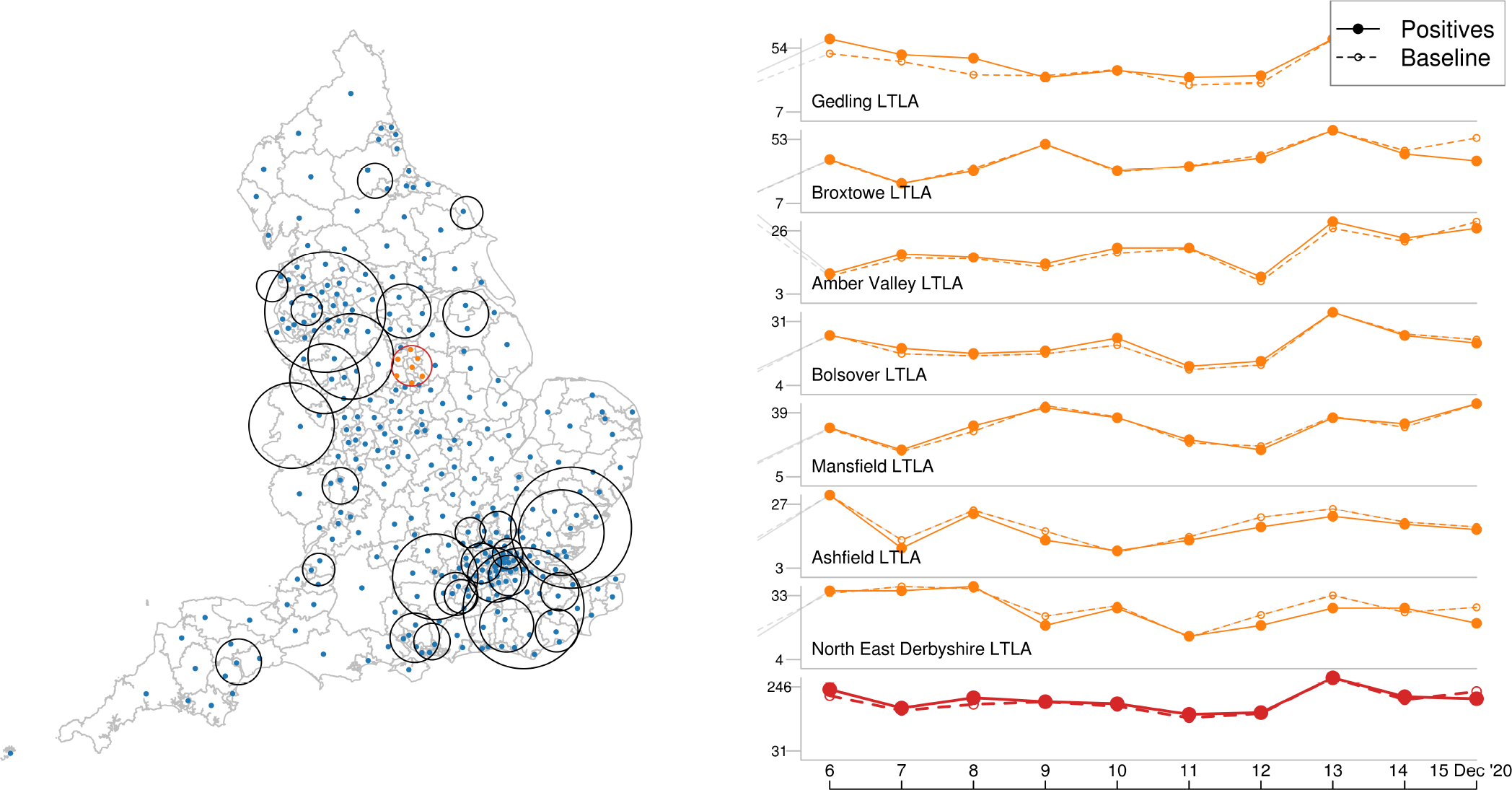
Illustration of RaNCover approach. Many overlapping cylinders with circular bases enclosing several LTLAs centres and heights corresponding to time intervals are drawn (left). For a generic cylinder *C* (e.g., the one with red circular basis), the observed number of cases from daily data in the enclosed LTLAs and the null-model intensities are aggregated (right, orange lines are positive test numbers in the LTLAs and red lines are total counts in the circle). In the highlighted cylinder, the total number of positive results is 1,809, while the baseline prediction is*≈* 1,957. According to criterion (5) the cylinder is not flagged.

It is not desirable to have a very low baseline, since this could trigger a signal even with one or two cases. We therefore heuristically perform the substitution 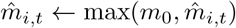to mitigate this effect. The cutoff *m*_0_ is chosen as the lowest value of intensity that does not trigger a warning if there is one S-gene positive case observed, with 𝒫 (*X >* 1 |*m*_0_) = 0.95. To evaluate the effect of decreased testing capacity, we considered a second scenario with half numbers of both S-gene positive and negative test results (rounded to the nearest integer for each time and location, see electronic supplementary text S3) and computed new warning scores using formula (6) with the new values of *M*_*i,t*_ and 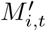. As a by-product of the procedure used to calculate the warning scores, we also obtain, for each cylinder, the fraction of positive (negative) tests over all test results in the area and time delimited by the cylinder. A moderated estimate of *M*_*i,t*_ (or 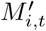) is the average of the ratio in all cylinders that contain (*i, t*). This generates a spatially and temporally averaged value for the proportion of all suitable tests that are S-gene positive (or S-gene negative) providing a measure of the invasion of new variants over time (see electronic supplementary material Movie S1).

## 3 Results

For each of the main four SARS-CoV-2 variants that emerged during the study period (Alpha, Delta, Omicron BA.1, and Omicron BA.2), we study patterns of S-gene test results and warning scores over time in all of England; aggregated test data are illustrated in figure 2, along with key dates of non-pharmaceutical interventions and VOC definitions as reported in [7, 32].

**Figure 2.**
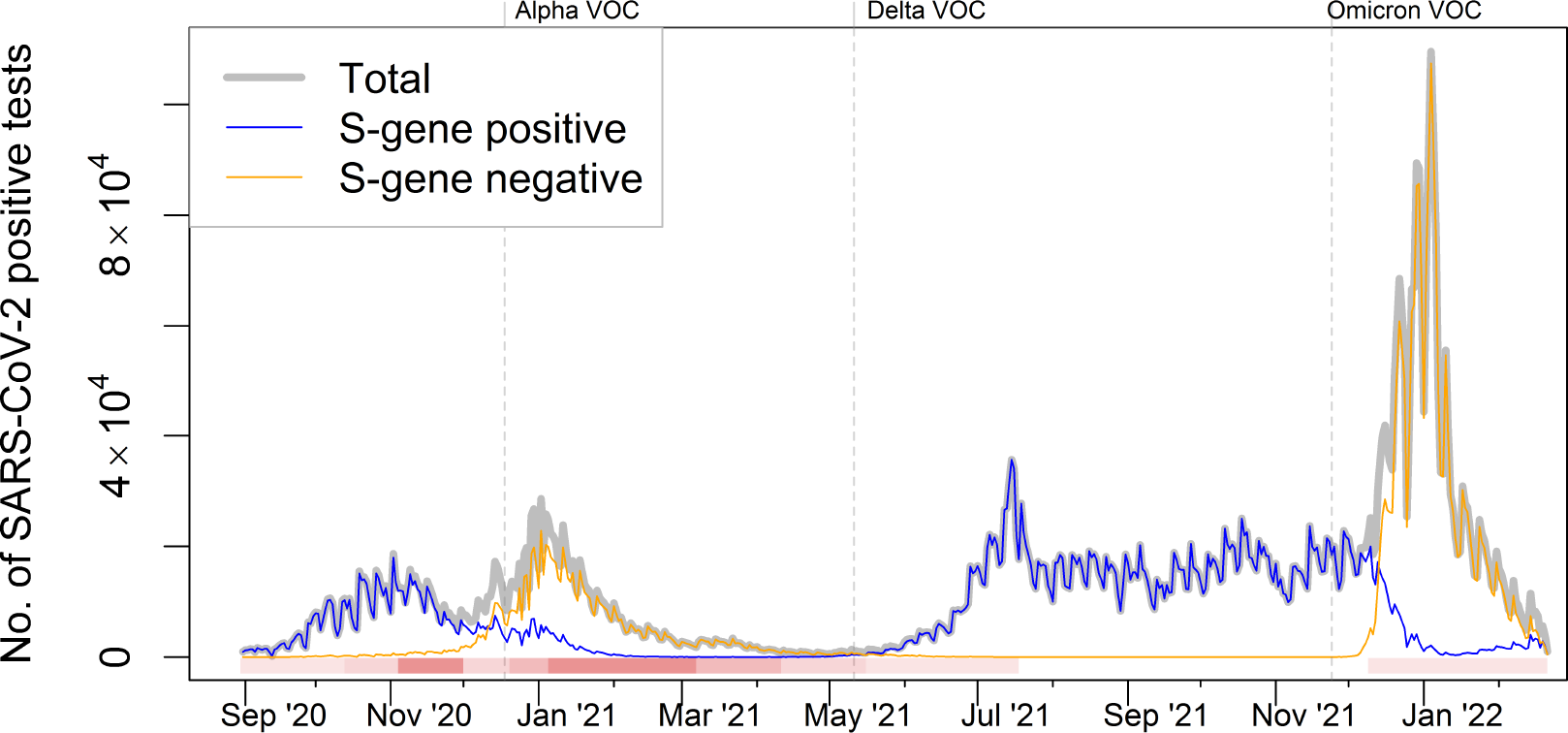
Pillar-2 test results reported in England over the study period. The solid grey line shows the total number of SARS-CoV-2 positive tests, which either tested negative or positive to S-gene target (blue or orange lines, respectively). This data set gives a proxy for the relative prevalence of a variant, with peaks corresponding to intensified testing or increased SARS-CoV-2 prevalence. Increased S-gene negative tests at the beginning of 2021 corresponds to Alpha invasion. The S-gene negative peaks in July 2021 is due to the Delta variant and the growth in S-gene negative tests at the end of 2021 corresponds to the first Omicron invasion. The horizontal bar highlights the introduction and relaxation of restriction measures (shades of red) on the following key dates: a) 14 October 2020, introduction of the 3-tier system to control local outbreaks; b) 5 November 2020, enforcement of the second national lockdown; c) 2 December 2020, lockdown ends and reintroduction of a (stricter) 3-tier system; d) 21 December 2020, introduction of the 4-tier system to control Alpha outbreak; e) 6 January 2021, the third national lockdown starts; f) 8 March, 12 April, 17 May, and 19 July 2021, 4-step roadmap to full relaxation of restrictions; g) 20 December 2021, mask mandate for public spaces to control Omicron. The three vertical dashed lines mark the dates when Alpha, Delta, and Omicron were declared Variants of Concern (VOCs).

### Alpha Variant

The SARS-CoV-2 variant of concern named B.1.1.7 (PANGO nomenclature [33]), also referred to as Alpha, has a specific mutation in the S-gene which results in a deletion of two amino acids of the spike protein (69-70del) and failure of S-gene target amplification in the TaqPath assay [34]. The proportion of S-gene target failures (SGTFs) over SARS-CoV-2 positive TaqPath PCR tests is thus a proxy for the relative prevalence of Alpha compared to other non-SGTF variants. Alpha was first detected in the COVID-19 Genomics UK Consortium (COG-UK, [35]) genome data in Kent on 20 September 2020 and spread quickly in London and then in other parts of England. Based on SGTFs, it is estimated that at the beginning of November 2020 5% of all cases were ascribed to this variant. Alpha was noted as a potential variant of concern and was designated a VOC on 18 December 2020 [7, 26], although the likely implications of this first (major) new variant were not yet fully understood on 2 December 2020, when the second lockdown period ended in England. By the end of January 2021, more than 95% of all new COVID-19 cases in England were likely caused by Alpha. While this VOC had greater transmissibility than its predecessors, its spread was driven by the increased movement of people across dominant source locations such as London [36]. As Alpha was first detected in South East England, and so effectively spread from a point location, our dataset provides an opportunity to test how quickly its potential as a VOC could have been assessed given epidemiological data from the variant’s emergence up until its designation as a VOC.

Some degree of geographic spread can be observed in data aggregated at regional level (solid lines in figure 3-A–B); indeed, by 19th November 2020 a quarter of all cases in the South East were attributable to Alpha, followed by reaching a quarter SGTF in the East of England (21 November), London (24 November), and South West (6 December) NHS regions. Aggregating data at such large scales hides local details, such as which Lower Tier Local Authorities (LTLAs) are invaded earlier than the others. However, with fewer tests available in a small region, detecting a local outbreak early can be difficult. This is therefore an ideal test of the RaNCover algorithm to highlight patterns on a finer spatial scale. A key Alpha hot spot appeared to be Folkestone and Hythe (an LTLA in the South East region of England) where the warning score first peaked (*w >* 90%) on 15 October 2020 (with only 5 SGFTs and 10 SARS-CoV-2 positive tests on that day) as illustrated in figures 3-C. In early November 2020, the warning scores increased in a large area including Greater London and surrounding LTLAs (see, e.g., figure 3-D), forewarning an outbreak in this area even though the fraction of detected S-negative tests in single LTLAs was still below 17% (lower than the levels reached in Folkestone and Hythe mid-October). The timeline of when each LTLA achieved a 90% warning score threshold, and hence a measure of the geographical spread of the Alpha variant is summarised in figure 4. During the initial phase of Alpha spread, anomalies were detected in different locations across England, presumably corresponding to S-negative variants (Alpha or others) which were not able to locally out-compete the wild-type (see electronic supplementary material Movie S1). Since 69-70del mutation also existed in other variants, these signals could also indicate variants which did not fix in the population. At the end of 2020 almost all LTLAs in England were flagged by high warning scores (figure 4); later, warning scores began to reduce as the presence of Alpha became the norm, starting from the most southern and eastern LTLAs, although some LTLAs in the North East (around Manchester) flagged up until the end of January 2021 (see electronic supplementary material Movie S1).

**Figure 3.**
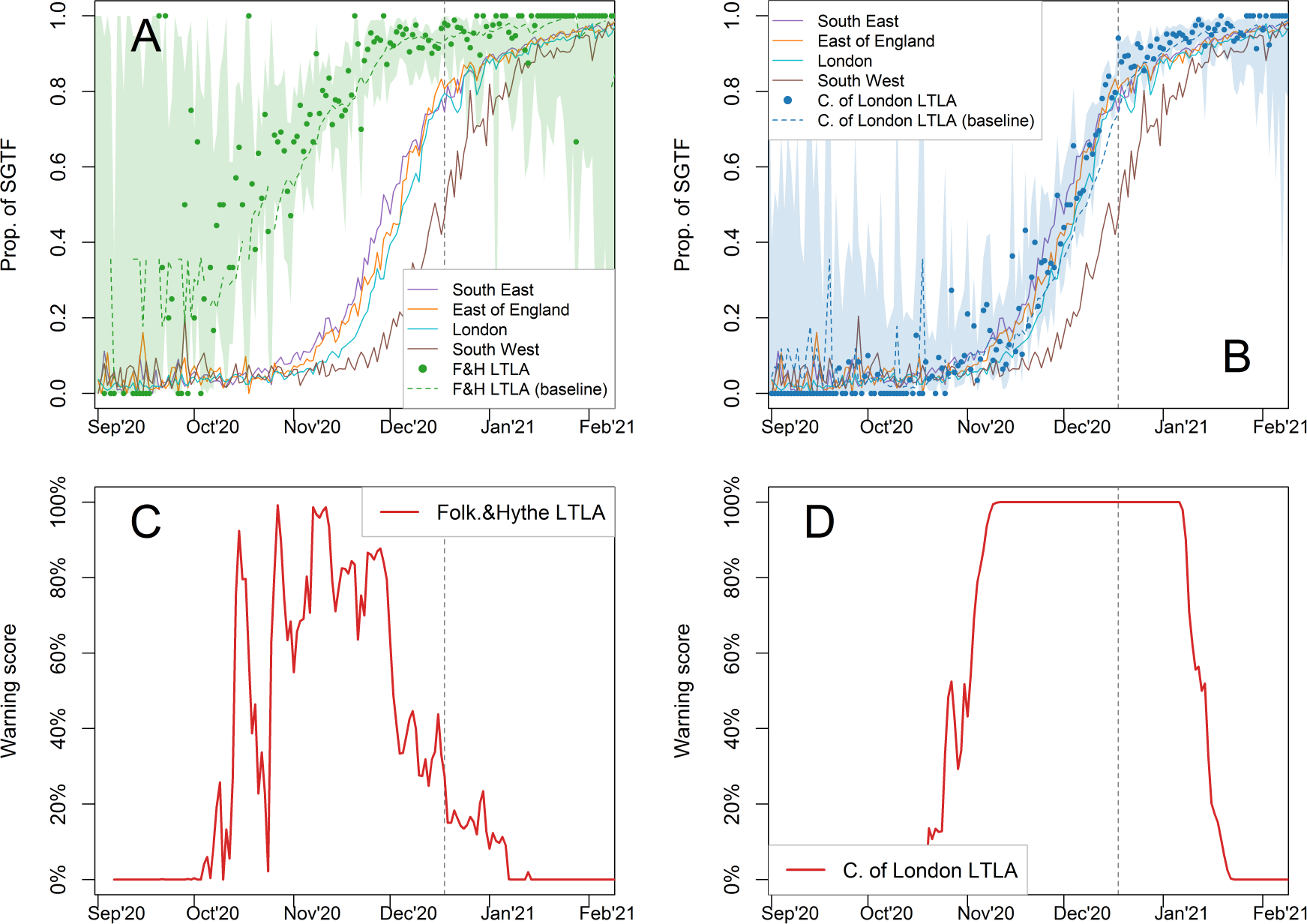
Progression of the Alpha variant in selected regions and LTLAs. A) Green dots are the proportion of SGTF in Folkestone and Hythe (F&H), with shaded areas representing 0.25%-97.5% Wilson CIs; uncertainly is substantial due to the low number of SARS-CoV-2 positive tests. For comparison, proportions of SGTFs per region average in South East, East of England, London, and South West are reported in green, magenta, blue, and grey solid lines, respectively. The green dashed line represents the expected proportion in F&H according to the null-model baseline. B) Proportion of SGTF tests in the City of London (blue dots) compared with local baseline (dashed blue line) and SGTFs proportions per region average in South East, East of England, London, and South West reported for comparison. C) RaNCover detects anomalies in F&H (red line is the warning score temporal trend) thus suggesting that the observed proportion of SGTF is significantly higher than the null model baseline. D) Increased warning scores (*w >* 90%) in City of London LTLA (red line) clearly flags the presence of an outbreak from 7 November 2020, even if the proportion of SGTF is still below the level reached in F&H. As in figure 2, the dashed vertical lines mark the Alpha VOC definition date.

**Figure 4.**
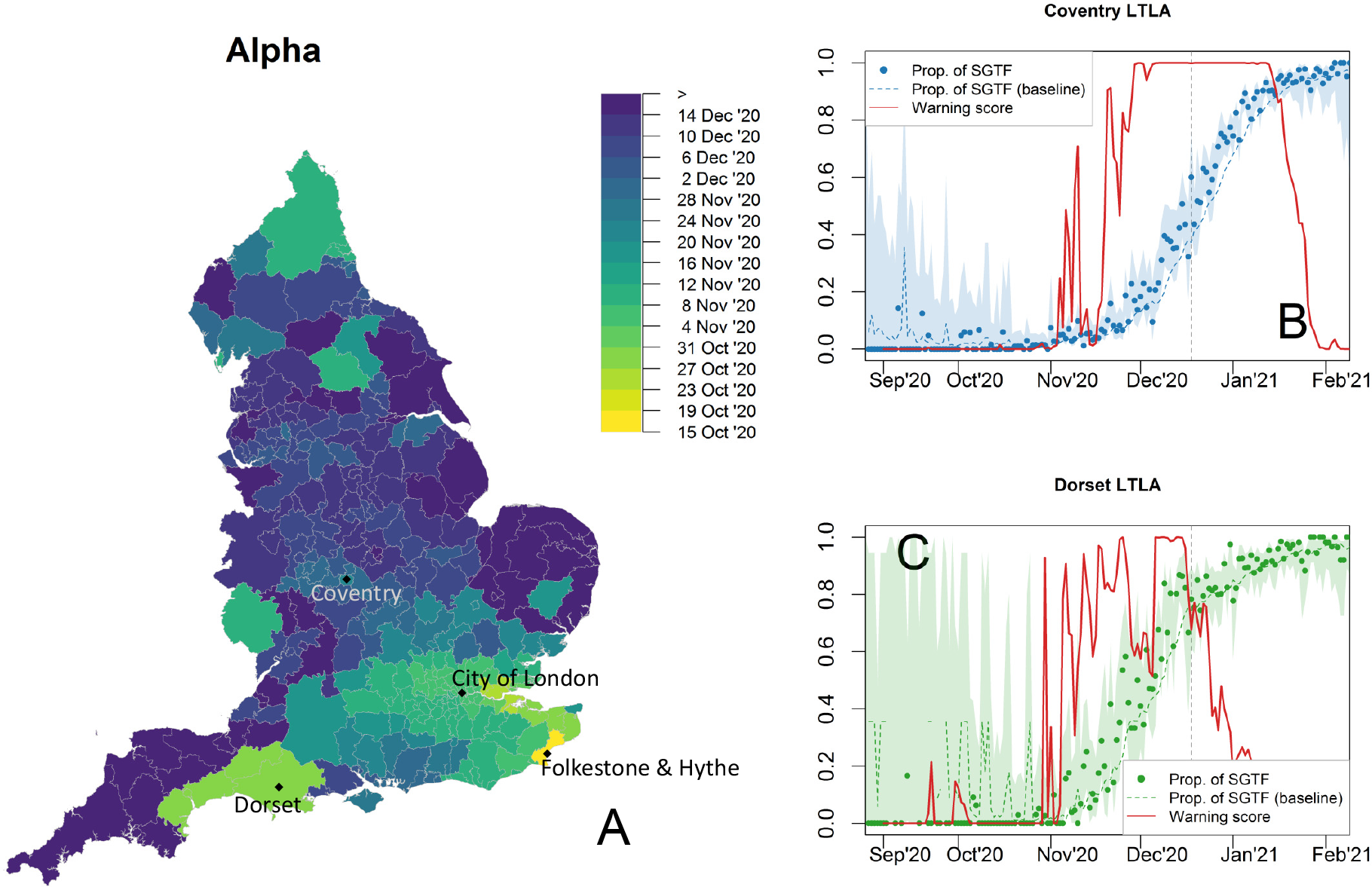
Progression of the Alpha variant and warning scores. A) Spatial diffusion of warning flags for SGTFs during the Alpha invasion. The colour map from yellow to blue indicates the date when an LTLA warning score first exceeded a threshold value of 90%, starting from 15 October 2020, when Folkestone and Hythe LTLA is first flagged (see figure 3). LTLAs in the South and Greater London are flagged first. Temporal trends in Coventry (B) and Dorset LTLAs (C). The observed proportions of SGTFs are marked by dots, with dashed lines representing null-model expectations and shaded areas 0.25%-97.5% Wilson CIs for proportions. Warning scores are represented by solid red lines. Vertical dashed lines mark the WHO declaration of Alpha as VOC on 11 May 2021.

The strength of the warning-score patterns depends on the total number of SARS-CoV-2 tests. With fewer tests, the proportion of SGFTs is estimated with wider confidence intervals and the increased uncertainty is also reflected in the warning scores; indeed the LTLAs in less populated regions, e.g., Folkestone and Hythe (see figure 3-C) and Dorset (figure 4-C), display fluctuating warning scores, especially at beginning of the outbreak. Nevertheless, warning flags are explicitly raised. Using down-sampled data (section 2.2) yields slightly delayed signals. Folkestone and Hythe exceeds the 90% threshold on 26 October 2020 (11 days later than the full data result and with 68% of SGFTs, but with only 16 SARS-CoV-2 positive tests). City of London is flagged only two days after the full sample date, with three SGFTs over a total of 26 SARS-CoV-2 tests (instead of three negatives over 18 tests from the full-sample warning date).

### Delta Variant

Starting from 6th January 2021 (when the third lockdown began), England experienced a period with no spread of novel variants of concern and decreased levels of SARS-CoV-2 positive tests. The VOCs that emerged directly after Alpha did not have the same spike-protein deletion as Alpha, thus testing S-gene positive in the TaqPath assay, and providing a means of rapid identification against a background of Alpha cases. The most common of these variants were B.1.351 (first identified in South Africa), P.1 (in Brazil), and B.1.617 PANGO lineages – with sublineage B.1.617.2 (also referred to as Delta) first identified in India, escalated in the UK on 6 May 2021 [27] and designed VOC by the WHO on 11 May 2021 [7]. S-gene positive results provided rapid signals for investigating community spread of these variants [21]. Phylogenetic studies demonstrated that Delta sublineages were introduced more than 1000 times from India before travel restrictions were introduced on 23 April 2021 [37]. When COVID-19 restrictions were relaxed on 17 May 2021 [38] (as England entered Step 3 of the relaxation process), all sublineages spread from independent seeding events and grew at a similar pace, with associated Delta clusters initially documented in the North West region [37]. Our data show that in England Delta overtook Alpha in less than three months, from May to June 2021. To detect local anomalies in S-gene positive test incidence we again used RaNCover. The North East area including and surrounding Greater Manchester was acknowledged at the time as a hotspot [39]; our warning score in this region first exceeded 90% in Warrington LTLA on 2 May 2021 (with Manchester, the most populated LTLA in the region, exceeding 90% on 7 May, figure 5-A). Interestingly, RaNCover flagged the South West LTLA of Somerset West and Taunton significantly earlier than elsewhere, with *w >* 90% on 23 April 2021. This suggests that an early outbreak of Delta (or another S-gene positive variant) might have occurred in that area unreported (figure 5-B and C). A London outbreak followed, with warning score exceeding 90% in the City of London LTLA on 19 May 2021.

**Figure 5.**
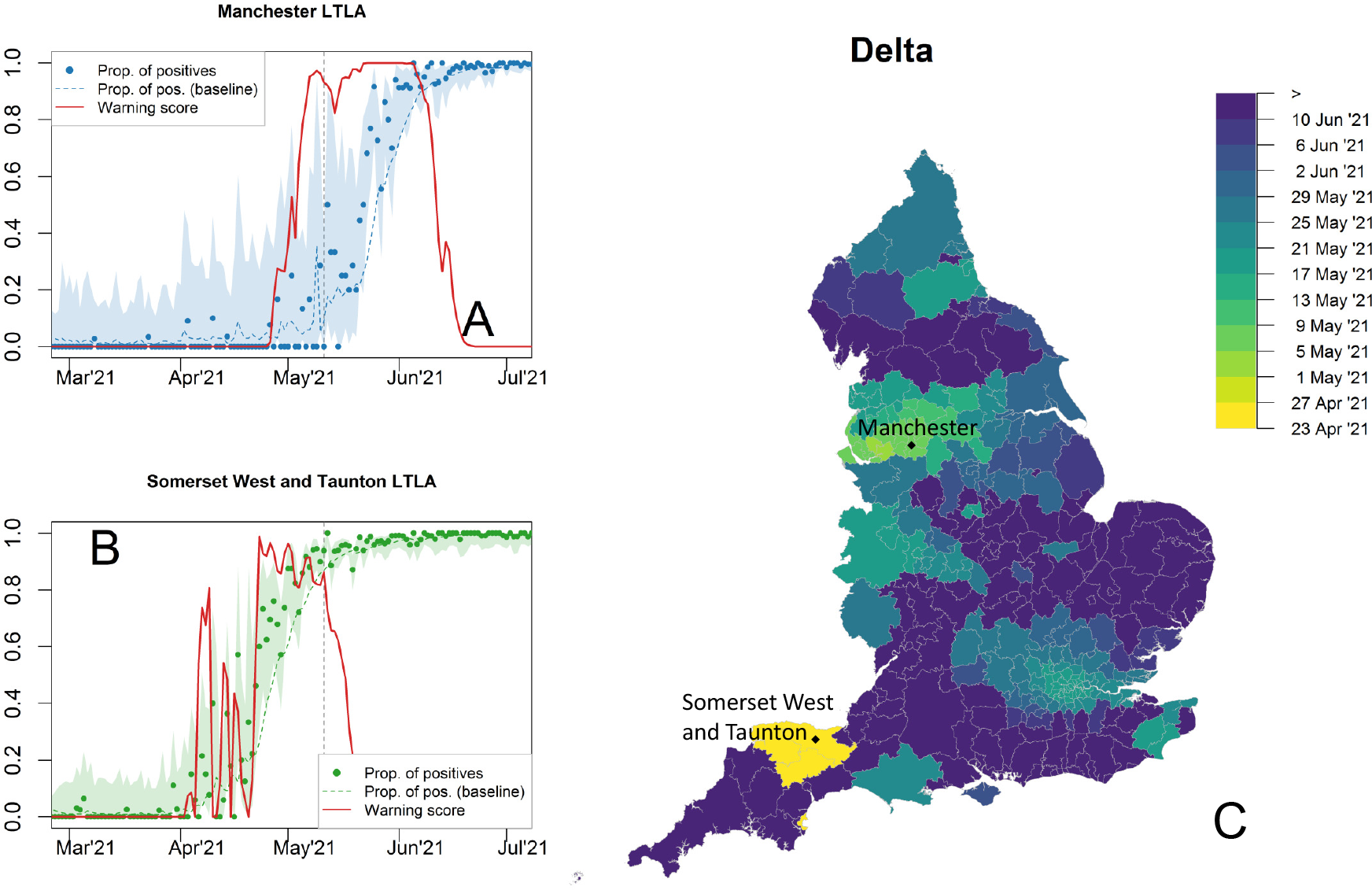
Progression of the Delta variant and warning scores for the proportion of S-gene positive tests. Temporal trends in Manchester (A) and Somerset-West-and-Taunton LTLAs (B). The observed proportions of tests are marked by dots, with dashed lines representing the expectations according to the null-model baseline. Shaded areas 0.25%-97.5% Wilson CIs for proportions. The warning score trend in Somerset West and Taunton LTLA flags the presence of aberrations from the null model, with *w >* 90%, on 23 April 2021. Manchester LTLA (North West region) has *w >* 90% on 7 May 2021; on that day the fraction of S-positive tests was reported as zero. The vertical lines mark the WHO declaration or Delta as VOC on 11 May 2021 (dashed line). C) Spatial diffusion of warning flags for S-gene positive tests during the Delta invasion. The colour map indicates the date when an LTLA warning score first exceeded a threshold value of 90%, starting from 23 April 2021. LTLAs in Somerset West and Taunton, East Devon, Mid Devon, and Torbay (yellow colour) are flagged first, followed by the North West (starting from Warrington LTLA) and London areas.

Delta invasion occurred in a period of decreased SARS-CoV-2 positive tests, as also illustrated in figure 2 (note the minima in the total number of positive tests at the beginning of May 2021). This obviously affects the quality of an early warning signal system. Using only half of the available data, warning scores did not reach the 90% level in certain LTLAs, yet they still reported the presence of anomalies. For example, in Warrington LTLA and Somerset West and Taunton LTLA, the warning scores peaked at *w* = 0.88% on 8 May and at *w* = 0.86% on 26 April 2021, respectively. In this scenario, Manchester and City of London LTLAs reached *w >* 90% respectively 12 and 5 days after the dates obtained from processing all available tests, thus demonstrating, on the one hand, the importance of widespread testing coverage; on the other hand, it is worth noting that RaNCover provides useful information even in this low-testing scenario (see figure S4 in supplementary text).

### Omicron

The Delta variant dominated in England throughout the summer and autumn of 2021. On 27 November 2021, the S-gene negative Omicron variant (BA.1) was first detected in the UK after being imported from South Africa. At the time of detection, it was deemed that the variant was likely to result in a rapid increase in cases due increased transmissibility and immune evasion and it was declared a VOC on 26 November 2021 [7, 28]. It was also observed that this VOC was associated with decreased severity [7, 40]. To slow down Omicron spread in England’s largely vaccinated population, a mask mandate was re-introduced on 20 December 2021. The Omicron wave in England was also characterised by a peaking number of SARS-CoV-2 positive tests (figure 2; this may be due to increased prevalence, increased testing capacity and increased awareness). The Omicron lineages have more than 30 mutations in the spike protein alone. In particular, the original Omicron variant, called BA.1, also shows 69-70del, thus resulting in SGTFs in the TaqPath tests. Omicron BA.2 has fewer mutations than BA.1 and lacks the genetic deletion on the spike protein that causes SGTFs; it has also been reported to be more transmissible than BA.1 [41, 42].

In England the S-gene negative Omicron (BA.1) variant spread during November and December 2021; overtaking Delta with more than 50% of all cases on 12 December 2021 and reaching 95% prevalence on 26 December 2021 according to the TaqPath testing data set. The LTLAs flagged first by the application of RaNCover are Bury, in the Manchester area (figure 6-A), and Northampton, in the Midlands, which exceeded a 90% warning level on 1 December 2021. This was a clear early warning, anticipating the Omicron wave, especially given that total number of BA.1 Omicron cases confirmed by genotyping across the whole England by this time was only 22 [43]. Due to the large number of tests during December 2021, the proportion of S-gene negative tests within an LTLA is estimated with high confidence, which yields strong early warning signals with RaNCover achieving *w >* 90% with only 0.03%, 0.26%, and 0.17% of SGTF in Bury, Northampton, and City of London LTLAs, respectively (figure 6-A, B, and C). In these three LTLAs the warning scores obtained from the down-sampled data set respectively exceeded the 90% threshold only four, one, and two days after the dates marked by the full data set, respectively (see also figure S5 in supplementary text). The short delay is ascribed to an exceptionally large number of SARS-CoV-2 positive tests during this phase of the epidemic (see figure 2).

**Figure 6.**
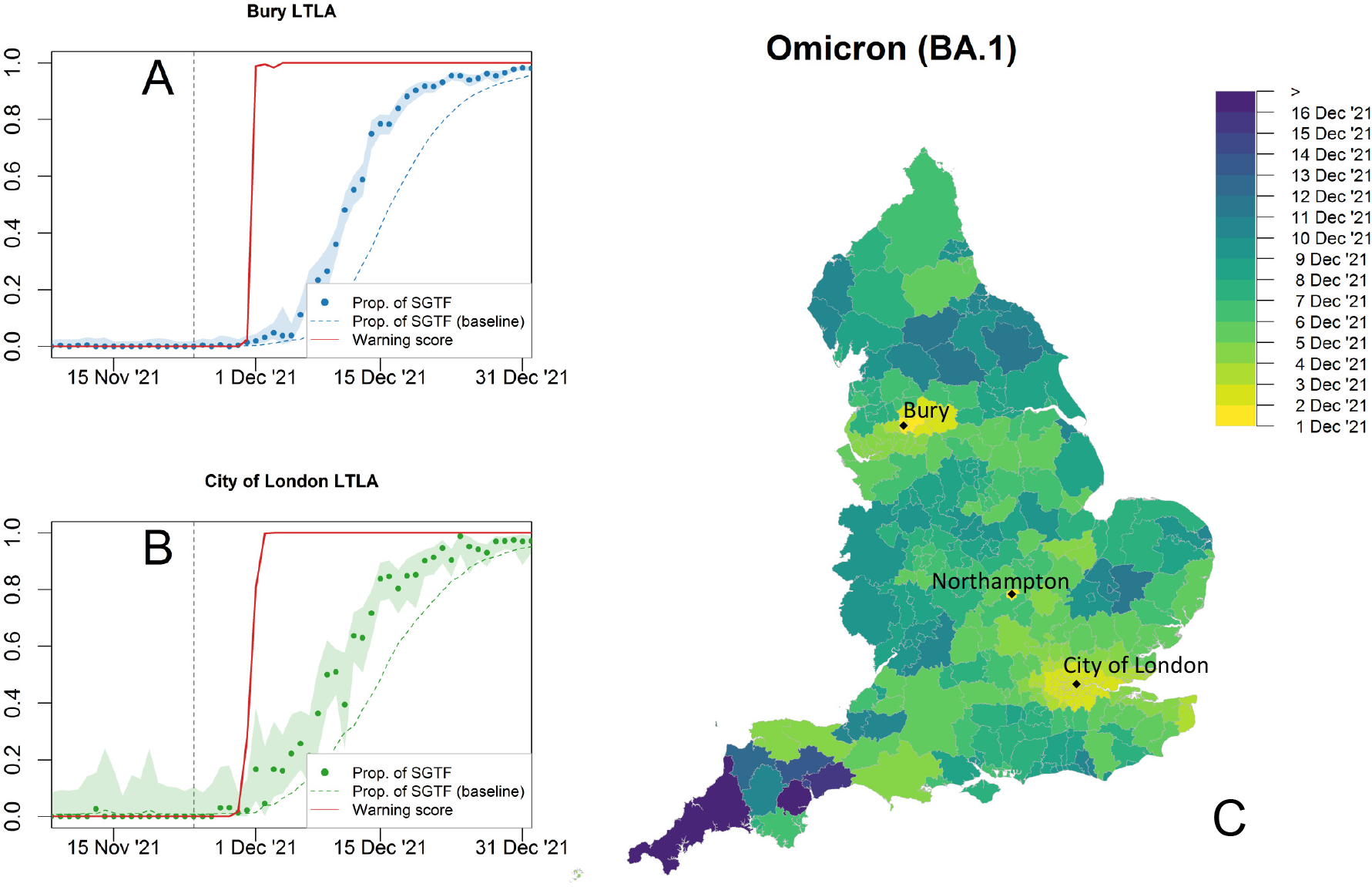
Progression of the S-gene negative Omicron variant (BA.1). Temporal trends and warning flag in Bury (A) and City of London (B) LTLAs. The observed proportions of tests are marked by dots, with dashed lines representing the expectations according to the null-model baseline. Shaded areas are 0.25%-97.5% Wilson CIs for proportions. The warning scores (solid red lines) display abrupt departure from the null-model expectations, with *w >* 90% on 2 December 2021 in the City of London and on 1 December 2021 in Bury and Northampton LTLAs. At those dates the reported fractions of SGTF in the single LTLAs were very low. The grey dashed vertical line marks the WHO declaration of Omicron as VOC on 26 November 2021 (see also figure 2). C) Spatial diffusion of warning flags during the Omicron BA.1 invasion. The colour map indicates the date when an LTLA warning score first exceeded a threshold value of 90% starting from 1 December 2021. The areas flagged first are Manchester, Northampton, and London.

A second Omicron invasion began in early 2022, with increasing numbers of S-gene positive (Omicron BA.2) cases in LTLAs in the London area, see figure 7. As of 10 January 2022, 53 sequences of the S-gene positive BA.2 sub-lineage of Omicron had been identified in the United Kingdom [43]. RaNCover raises a warning signal in the City of London with *w >* 90% on 17 January 2022 (18 January using the down-sampled data), when only the 0.01% of positive SARS-CoV-2 tests in that LTLA were positive to the S-gene target (figure 7-A). By the end of the study period on 20 February 2022, the Omicron BA.2 variant was set to replace the S-gene negative Omicron.

**Figure 7.**
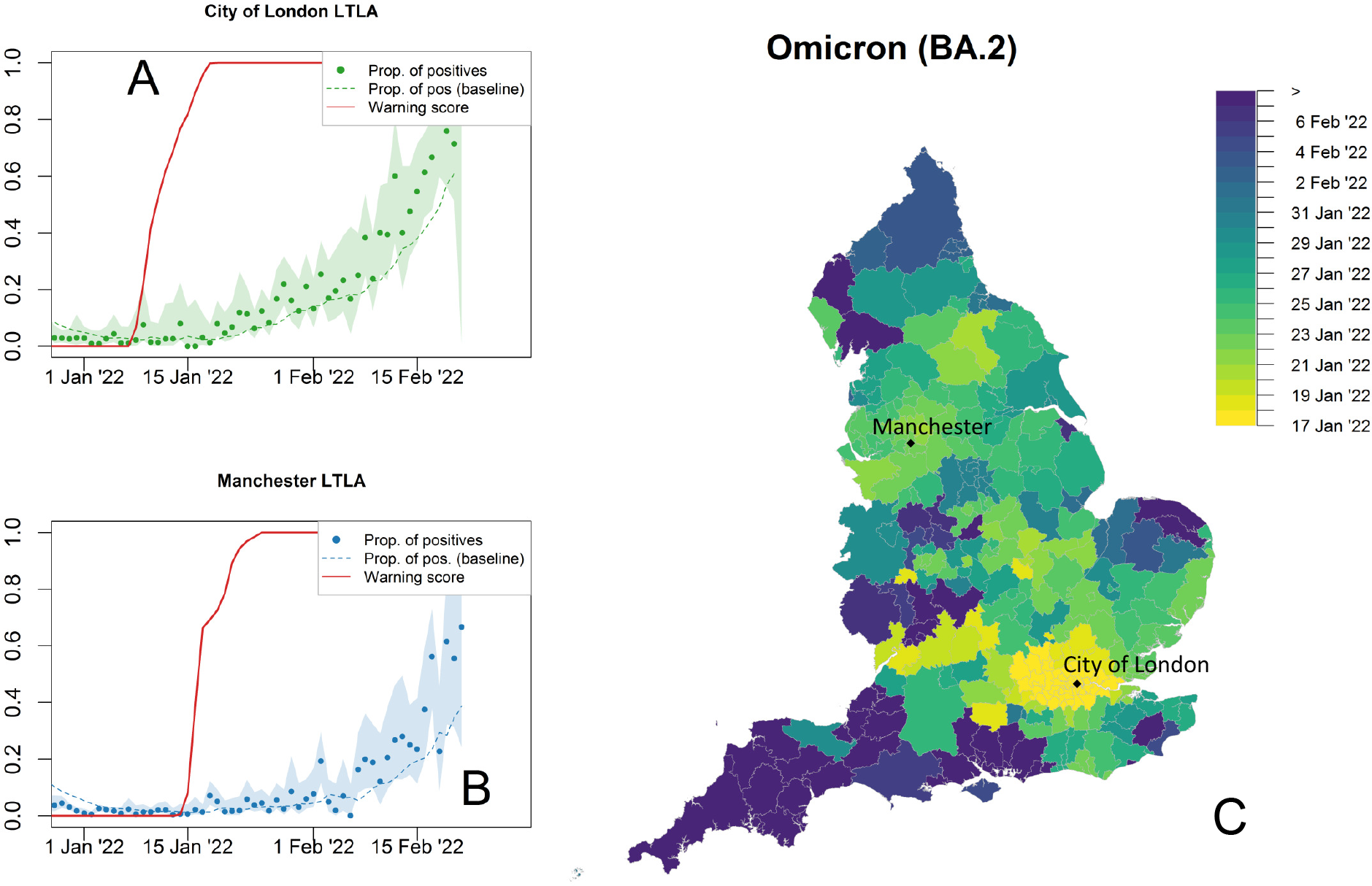
Progression of the S-gene positive Omicron variant (BA.2). Temporal trends in the City of London (A) and Manchester (B) LTLAs. The warning scores reach the threshold value (*w >* 90%) on 17 January 2022 (City of London) and on 23 January 2022 (Manchester), despite the fraction of S-gene positive tests being still very low (0.01 and 0.04, respectively). Shaded areas are 0.25%-97.5% Wilson CIs for proportions. C) Spatial diffusion of warning flags for SCGF during the Omicron BA.2 invasion. The colour map indicates the date when an LTLA warning score first exceeded a threshold value of 90%, starting from 17 January 2022.

### Local anomalies of the dominant variant

So far we have generated warning scores by considering statistically significant increases in the non-dominant S-gene type (e.g., increases in the proportion of S-gene negative tests when the dominant variant was S-gene positive). This provided a signal for invasion of a new variant (assuming it is different in terms of S-gene detection). However, our methodology can also be applied to anomalies in the S-gene type of the dominant variant, due to increases above the expected baseline which never achieves 100% dominance (figures S7 and S8 in supplementary text). As an example, we initially consider the proportion of S-gene positive tests during October 2020 (figure S7 A and D in supplementary text), when we observe wide-spread and diffuse warning scores due to an increase in the proportion of S-gene positive samples generating multiple exceeding warning scores (*w >* 90%). In the absence of detailed genomic investigation, it is difficult to state whether these correspond to local outbreaks in wild-type SARS-CoV-2 (or other non-documented S-gene positive variants) or to random fluctuations in the early testing regimen. In early October 2020, we also observed a transient warning score for S-gene negative variants in the North West of England (figure S7 C in supplementary text). This is the only clear false positive signal of variant change observed in our study. It is again unclear of the reasons behind this signal, it could be due to a short-lived invasion of a S-gene negative variant, it could be a consequence of higher than normal levels of S-gene positive tests in the rest of the country (as illustrated by figure S7 A in supplementary text), or it could be a random fluctuation due to low levels of testing at this time. Patterns for local variations in the dominant S-gene type can observed throughout 2021. In March 2021, during the Alpha wave, there are raised warning scores (*w >* 90%) in East Anglia with an increase in the dominant S-gene negative tests (figure S8 A-B in supplementary text). During the Delta wave there are high warning scores due to an increase in the proportion of S-gene positive tests (figure S8 C-D in supplementary text), clustered in four separate occasions associated with periods of increase in testing.

## 4 Discussion

Here we have used data from community PCR testing for SARS-CoV-2 generated by the TaqPath system [20, 21] to explore the early invasion of new variants of concern. The TaqPath system was designed to accurately diagnose SARS-CoV-2 infections. It is also able to detect the S-gene present in the ancestral wildtype variant, the Delta variant, and the Omicron BA.2 variant, while it fails to amplify the S-gene target in Alpha and Omicron BA.1 variants, thus providing a way to discern if the infection is due to one of these variants. In fact, the system has been used as a rapid and reliable means of tracking new variants [20–22, 24, 36], which operated more rapidly and at a greater scale than genomics testing [44]. The fortuitous pattern of invasion of SARS-CoV-2 variants with alternating S-gene target results, spanning over one year from September 2020 up until February 2022, also provided an ideal avenue for studying the spread of SARS-CoV-2 variants and evaluating outbreak detection methodologies. The growth and detection of new variants is generally subject to considerable stochastic fluctuations; as such, simple methods of estimating the early growth rate or looking for changes are error-prone in the early stage of invasion. Simple methods may also be influenced by changes in testing or contact behaviour, which will affect the observed growth of both resident and invading variants. Here we use the RaNCover methodology, previously developed to detect anomalies in disease patterns subject to strong fluctuations due to low count numbers (with the initial case study focused on the detection of invasive group A Streptococcus outbreaks [16]). We apply this to the proportion of positive cases which are S-gene negative (for invasion of Alpha and Omicron BA.1) or positive (for Delta and Omicron BA.2), thereby reducing the impact of changes in testing behaviour.

RaNCover is a method to highlight aberrations from a baseline level that uses as its null model the observations expected in an idealised, steady-state, situation. The general approach consists of drawing, inspecting, and aggregating the information in a large number of spatio-temporal cylinders. In the original formulation of RaNCover [16], which was tailored to detect clustered outbreaks in sparsely distributed cases of infection, our null model assumed cases occurred at a Poisson rate proportional to population sizes in each location. Here we used a different baseline (null model) to study the relative proportion of SARS-CoV-2 variants in lower-tier local authorities (LTLAs) and detect early the invasion of a novel variant. We defined the baseline as the expected number of S-gene positive or negative tests (indicating the presence of a variant) under the assumption that the proportion of S-gene positive (or negative) samples detected is stationary. Therefore, an aberration from this baseline is an early warning signal that a new variant is increasing (relative to the resident variant) in a geographical area and that a local outbreak might occur. As this approach detects increases in proportion relative to other SARS-CoV-2 sublineages, it is relatively unaffected by changes in the overall number of detected cases (such as may be driven by variations in testing behaviour, social mixing or changes in population immunity). As a second step, we process the information in these cylinders and obtain a continuous warning score *w* (the larger the value of *w* the higher the warning level) for each location.

In this retrospective application to S-gene tests, we were able to rapidly detect each new variant as its prevalence started to increase within local populations. We detect signals of the rise of the Alpha variant in Folkestone and Hythe LTLA (in the South East of England) on 15 October 2020, nearly two months before it was declared a variant of concern. We detect early warning signals for the Delta variant from the area around Manchester on 6th May 2021, as well as for an earlier S-gene positive anomaly (potentially also due to Delta) around Somerset on 23 April 2021 which, to the best of our knowledge, has not been documented before. Again, these signals pre-date 11 May 2021, when the Delta variant was first declared a variant of concern. Given the large amount of testing in late 2021 and early 2022, we are able to rapidly detect the relative growth of Omicron variants (as shown by the tighter confidence intervals in figures 6 and 7 than in figure 3). Warning signals for Omicron BA.1 are detected in the North East and Greater London on 1 December 2021 and 2 December 2021 respectively, when the prevalence detected by other means was still low. Finally, the Omicron BA.2 variant generates a warning in Greater London on 17th January 2022. We only find one example of a potential false positive from the RaNCover methodology, in October 2020 when a slight increase in S-gene negative tests was detected in the North West of England centred around Kirklees, Oldham and Rochdale (figure S7 C in supplementary text). It is impossible to know whether this warning is a statistical fluctuation or a short-lived invasion without a detailed genomic investigation of cases over that time period.

These detection events, based on S-gene tests, benefit from hindsight. In October 2020, we were not expecting to observe new variants, and any changes to S-gene detection would not have been an immediate cause for alarm. We therefore recognise that detailed genomic investigation is needed to interpret data on circulating SARS-CoV-2 variants. PCR testing in England declined from its maximum of over a hundred thousand detected cases per day in early 2022 (when over half a million tests were performed each day), to under a thousand a day since 2nd April 2022 and continued to decline. This drop corresponded to the change in testing policy [45], when symptomatic testing was restricted to vulnerable and at-risk populations and extensive PCR-based testing was abandoned in favour of integrated genomic surveillance, as per WHO recommendations [2, 46].

Genomic sequencing remains the gold-standard method to identify which variant of SARS-CoV-2 is present in a specimen. A range of different Omicron variants have been found in genomically sequenced specimens after the end of the Omicron BA.2 outbreak in May 2022, denoted BA.4, BA.5, and BA.2.12.1, with BA.5 becoming dominant around August 2022 in the United Kingdom. At the time of writing, a number of additional variants, i.e., BA.2.75, BQ.1, XBB, XBB.1.5, and so-called recombinant variants (created by the combination of genetic material from two different variants), are simultaneously circulating [43, 47], thus demonstrating increasingly complex infectious disease dynamics, arguably resulting from interactions between multiple variants, immune escape, vaccination and other public health interventions, and behavioural changes. The presence of multiple variants also invalidates identification methods based on S-gene target only. However, the RaNCover approach presented here can be easily adapted to more comprehensive genomic data. In fact, genomic sequencing can be used to determine the precise viral variant (or sub-variant) that caused an infection case and this information can be aggregated to estimate proportion of each variant among all the sequenced samples at a particular time (as required by our methodology, see subsection 2.1), although this would introduce additional delays into the system and would reduce the accuracy of statistical approaches as the number of samples would likely be lower. Genomic surveillance has also the limitation that the observed frequency of variants may not be representative of population variant frequencies, mainly due to biased selection of samples sent for sequencing, especially after April 2022. However, effective surveillance does not require the sequencing of a specimen from every COVID-19 case, or even from random representative samples, but may perform well using just selected samples, e.g., patients in hospitals [48]. In this respect, our formulation of the baseline, conditioned on the number of tests per area, remains helpful and can be arguably extended to genomic surveillance contexts.

We reported that the generation of early warning signals greatly benefits from increased volumes of data generated by the large number of positive PCR tests obtained during the SARS-CoV-2 pandemic. As disease surveillance is rapidly shifting towards utilising genomics data, which permit simultaneous monitoring of more variants but have lower throughput than PCR, meticulous validation studies of early warning systems based on these sparser sources of data are required. For comparison, the application of genomic surveillance of SARS-CoV-2 in England has produced on average more than 38,000 sequenced SARS-CoV-2 samples per month, from January 2022 to February 2023, according to the data available at [49] – this is much less than the average *≈* 420,000 community cases per month detected with PCR during our study period. On the other hand, it is also worth noting that we encountered and studied a low-count scenario at the beginning of the Delta wave (see figure 2, with less than 1,000 positive PCR tests per day at the end of April 2021) and RaNCover was still able to get early warning signals during this phase, thus suggesting the wider applicability of our approach in genomic surveillance. As also discussed in reference [16], RaNCover is actually well suited to extract warning signals from sparse data.

One interesting spatial pattern that can be pulled from the continuous warning scores is the timing at which warning flags (e.g., with *w >* 90%) are first raised, before a variant predominantly spreads. Comparing figures 4 to 7 several factors in common emerge. Firstly, London is usually one of the first areas to generate flags; even for the Alpha and Delta variant that were initially centred elsewhere, London was the second and third geographic hotspot to be identified, respectively. This is presumably due to the number of transport links to London helping to facilitate the spread of infection, and the amount of international travel associated with the city bringing imports of novel variants. Secondly, there is generally a wave-like spread of warnings with the occasional long-range jumps, as would be expected from a spatial transmission kernel with a heavy tail [50]. Finally, the South West and to a lesser extent East Anglia are generally the last to experience warnings, which can be attributed to their lack of major cities and long journey times which mitigate the spread of infection. Data on the spatial spread of warning scores over time (as exemplified by figures 4 to 7) may be helpful to predict the likely time-scales for any new variant to spread before it becomes dominant, with Alpha and Delta taking around two months and Omicron BA.1 and BA.2 only requiring about two weeks (likely due to higher transmission and immune evasion of the variants and fewer public health interventions at the time).

The ability to rapidly detect the relative growth of a new variant in a region also has important public health implications. Any information on the heterogeneous distribution of infection allows resources to be targeted if necessary, helps inform local public health messaging and awareness or allows authorities to prepare local health services for increased burden. In England for example, a program of spatial targeting of controls (known as Tiers) was implemented from 14 October to 15 November 2020 and from 2 December 2020 to 6 January 2021 (see also figure 2); during this period different areas of the country were evaluated as being at “medium”, “high”, or “very high” alert levels, with different restrictions imposed depending on the severity of the outbreak and allocations regularly reviewed by the Joint Biostatistics Centre. From 21 December 2020, England also included a fourth “stay at home” tier, imposing additional restrictions to people’s movements in high-risk local authorities [32, 51]. More generally, by means of early warning signals specific to precise geographic locations, public health officials can quickly take action to contain or slow down a disease and reduce its overall impact if necessary.

One potential limitation of this study is that, in the absence of a ground truth which determines the scale of epidemic and the component variants with high certainty, it is difficult to quantify the accuracy of our results. Such comprehensive validation studies are often required for deployment of new methodologies (such as RaNCover) in real-world routine surveillance [52]. However we have already shown and discussed the accuracy of RaNCover’s accuracy in simulation experiments [16], and its performance with COVID-19 data adds extra support to our claims of accuracy and robustness. In fact, RaNCover’s approach offers an improvement from the usual viewpoint of monitoring and catching exceedances in individual locations, which are subject to important fluctuations that can cause false positive and false negative detection events. With its ability to aggregate signal from multiple regions, resulting in earlier identification of areas of concern, where a new variant is set to grow before it becomes widespread, we believe the random neighbourhood covering approach may be a valuable tool in the efforts to mitigate the impact of emerging infectious diseases.

## Data Availability

SARS-CoV-2 data were supplied under strict data protection protocols agreed between the researchers and UK Health Protection Agency. Interested readers wanting similar data may wish to contact, as detailed at https://www.gov.uk/government/publications/accessing-ukhsa-protected-data/accessing-ukhsa-protected-data. Spatial maps created with Sf using digital vector boundaries available from the Office for National Statistics, licensed under Open Government Licence v.3.0. Crown copyright and database right 2022. Terms and conditions of supply for the digital boundaries and reference maps are provided at https://www.ons.gov.uk/methodology/geography/licences. The scripts utilised in this project can be found at the following repository https://github.com/mcavallaro/SARS-CoV-2-spatiotemporal-surveillance.

## Supplementary material

[Movie S1](https://github.com/mcavallaro/SARS-CoV-2-spatiotemporal-surveillance/tree/master/Movie)

Supplementary text, sections S1, S2, S3, S4, and S5.

## Acknowledgements

The authors would like to thank Andre Charlett and the data teams at UKHSA for collecting and making available the data used in this study.

## Funding

This work was supported by Health Data Research UK, which is funded by the UK Medical Research Council, EPSRC, Economic and Social Research Council, Department of Health and Social Care (England), Chief Scientist Office of the Scottish Government Health and Social Care Directorates, Health and Social Care Research and Development Division (Welsh Government), Public Health Agency (Northern Ireland), British Heart Foundation and the Wellcome Trust (MC, MJK). MJK was also supported by the JUNIPER modelling consortium [grant number MR/V038613/1] and by the National Institute for Health Research (NIHR) [Policy Research Programme, Mathematical and Economic Modelling for Vaccination and Immunisation Evaluation, and Emergency Response; NIHR200411]. MJK is affiliated to the National Institute for Health Research Health Protection Research Units (NIHR HPRUs) in Gastrointestinal Infections and in Genomics and Enabling Data. This research is supported by the National Institute for Health and Care Research (NIHR) Applied Research Collaboration (ARC) West Midlands (DT). The views expressed are those of the author(s) and not necessarily those of the NIHR, the Department of Health and Social Care or Public Health England. The funders had no role in study design, data collection and analysis, decision to publish, or preparation of the manuscript.

## Data Availability

SARS-CoV-2 data were supplied under strict data protection protocols agreed between the researchers and UK Health Protection Agency. Interested readers wanting similar data may wish to contact, as detailed at https://www.gov.uk/government/publications/accessing-ukhsa-protected-data/accessing-ukhsa-protected-data. Spatial maps created with Sf [53] using digital vector boundaries available from the Office for National Statistics, licensed under Open Government Licence v.3.0. Crown copyright and database right 2022. Terms and conditions of supply for the digital boundaries and reference maps are provided at https://www.ons.gov.uk/methodology/geography/licences. The scripts utilised in this project can be found at the following repository https://github.com/mcavallaro/SARS-CoV-2-spatiotemporal-surveillance.

## Ethics statement

Data were supplied after anonymisation under strict data protection protocols agreed between the University of Warwick and UK Health Protection Agency, with public data deposition non-permissible for privacy reasons. The ethics of the use of these data for these purposes was agreed by UK Health Protection Agency with the Government’s SPI-M-O / SAGE committees.

## Supplementary text

### S1 Detection threshold

The detection threshold is the quantile function evaluated at 1− *α*. For discrete random variables, this is piecewise defined, It can be obtained for a Poisson random variable, e.g., with R using the built-in function `qpois(0.05, lambda, lower.tail=F)` (for a given value of the baseline represented by `lambda`). This function returns a discrete number which can be compared with the observation *M* according to expression (3).

In fact, the above is equivalent to computing the upper-tail cumulative probability for the Poisson random variable (which is ∈ ℛ) evaluated at *M*_*i,t*_ (`ppois(m[i,t], lambda, lower.tail=F)`) and compare with the threshold value *α* (exceedance occurs when the cumulative probability is lower than the chosen *α*.

### S2 A beta-binomial null model

The number *X*_*i,t*_ of positive results detected in (*i, t*) among *N*_*i,t*_ tests is represented by a binomial random variable, whose Poisson model of (2) is a limiting case for *N*_*i,t*_ → ∞, *p*_*i,t*_ → 0 and their product held finite, i.e.,

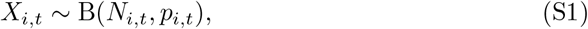

where the parameter *p*_*i,t*_ is the probability of testing positive in (*i, t*). We now set the probability *p*_*i,t*_ as another random variable that generates a fixed average proportion of 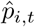 successes and has probability distribution *f* (*p*_*i,t*_|*N*_*i,t*_). For a given *p*_*i,t*_, the probability of 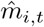successes in *N*_*i,t*_ tests is 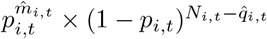.Assuming a Jeffrey beta prior distribution for it, i.e., 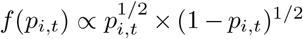 we get, by Bayes rule, that the probability distribution of *p*_*i,t*_ conditioned on *N*_*i,t*_ is also beta, with

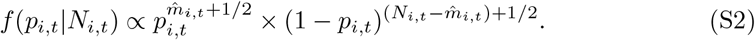

The number of tests *N*_*i,t*_ controls the dispersion of *p*_*i,t*_, with its distribution concentrating around 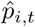 as *N*_*i,t*_ increases. This gives rise to a beta-binomial model for *X*_*i,t*_, which depends on the expected proportion 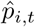 through equation (S1) and

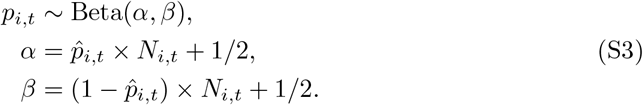

The likelihood of this model evaluated on our data set is higher than the Poisson model of the same mean (log-likelihoods -265,534.7 vs -410,335.4 and -260,616.4 vs -280,208.2 for the S-gene positive and negative case, respectively), suggesting that that model can be a valid first approximation. A model with fitted dispersion – *N*_*i,t*_→ *a* + *N*_*i,t*_*× b* in equation (S3), parameters *a* and *b* obtained by maximum-likelihood estimation – yields similar results. It can also be used analogously to criterion (3) to raise warning flags when the number of observed cases in a LTLA exceeds a chosen *α*-quantile threshold. However, this choice does not have a straightforward exceedance criterion for cylinders such as that of (5), which allow us to aggregate data from different cylinders and define the RaNCover warning scores. In figure S1 we compare the warning scores with the cumulative probability of obtaining a given count from the Poisson and beta-binomial models in selected LTLAs. The warning scores provide the clearest signal of the three. For 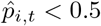, the beta-binomial distribution of (S1) and (S3) is over-dispersed with respect to the Poisson of (2) (figure S2). Therefore it is less likely to report exceedances during the initial phase of an outbreak.

**Figure S1.**
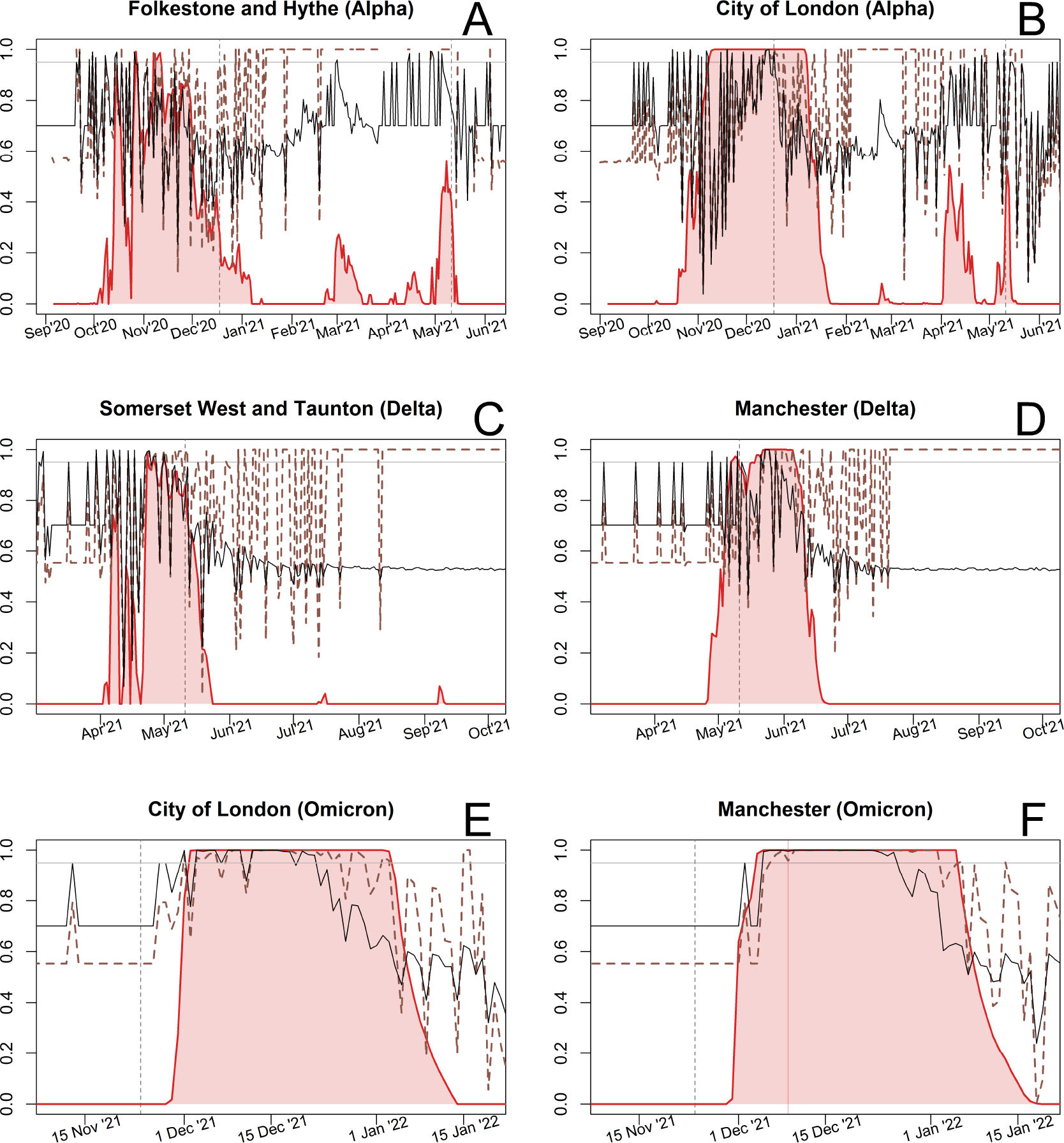
Comparison between RaNCover warning scores (red solid lines) and naive exceedance score based on single lower-tier local authorities (LTLAs). Naive scores are defined as cumulative probabilities of observed counts under Poisson (black lines) and beta-binomial null-models (brown dashed lines), and are highest when the observations lie in the upper tail of the null-model distribution. These values are subject to strong fluctuations due to low counts in single LTLAs. By incorporating information from neighbouring LTLAs, RaNCover highlights when a new variant is taking over more clearly than the single LTLA strategies.

**Figure S2.**
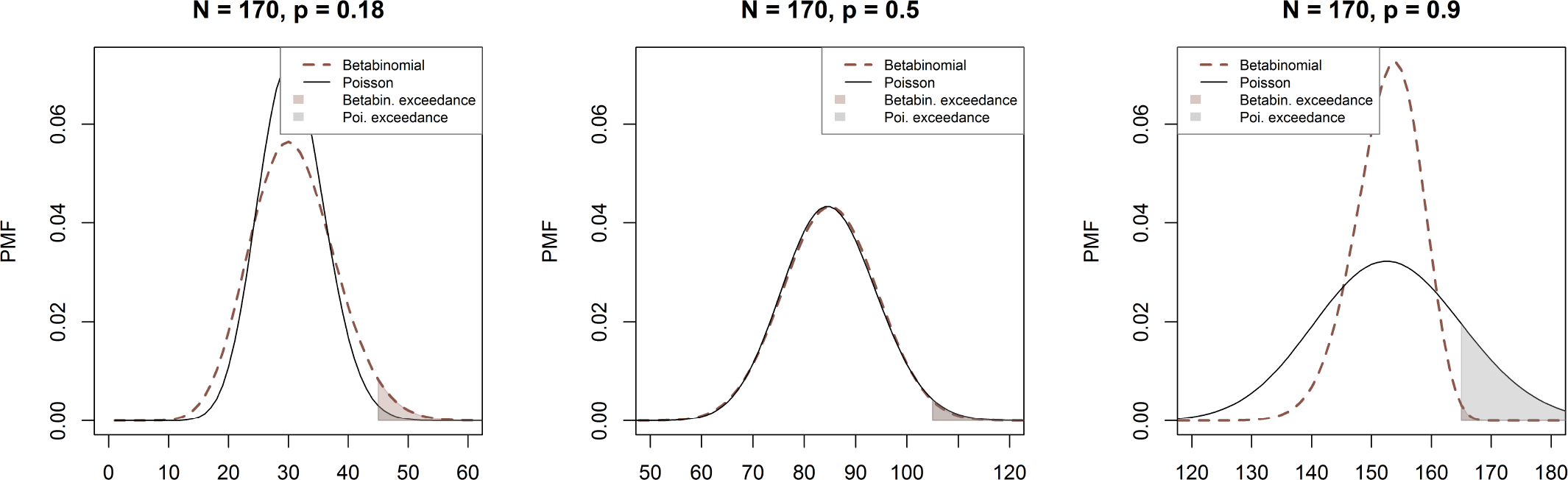
Comparison between Poisson and beta-binomial models of mean *p ×N* . Exceedance probabilities correspond to the shaded areas. When the parameter *p* is smaller than 0.5, the beta-binomial is over-dispersed and yields higher exceedance probability than the Poisson model (left plot). When *p >* 0.5, the beta-binomial is under-dispersed and its exceedance probability is lower than Poisson (right).

### S3 Down-sampled data results

To evaluate the effect of decreased testing capacity, we halved the numbers *M*_*i,t*_ and 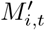 of S-gene positive and negative test results and rounded to the nearest integer,

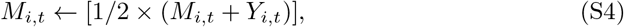

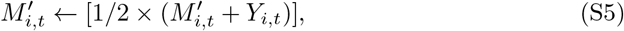

where *Y*_*i,t*_ is randomly chosen in *{*−0.1, 0.1*}*. We then computed the warning scores using the procedure detailed above, with 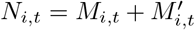 for each location and time *i, t*.

The warning results obtained with this down-sampled data set for the Alpha, Delta, Omicron BA.1, and Omicron BA.2 invasions are summarised in figures S3, S4, S5, and S6, respectively, which are analogous to figures 4, 5, 6, and 7 in the main text.

**Figure S3.**
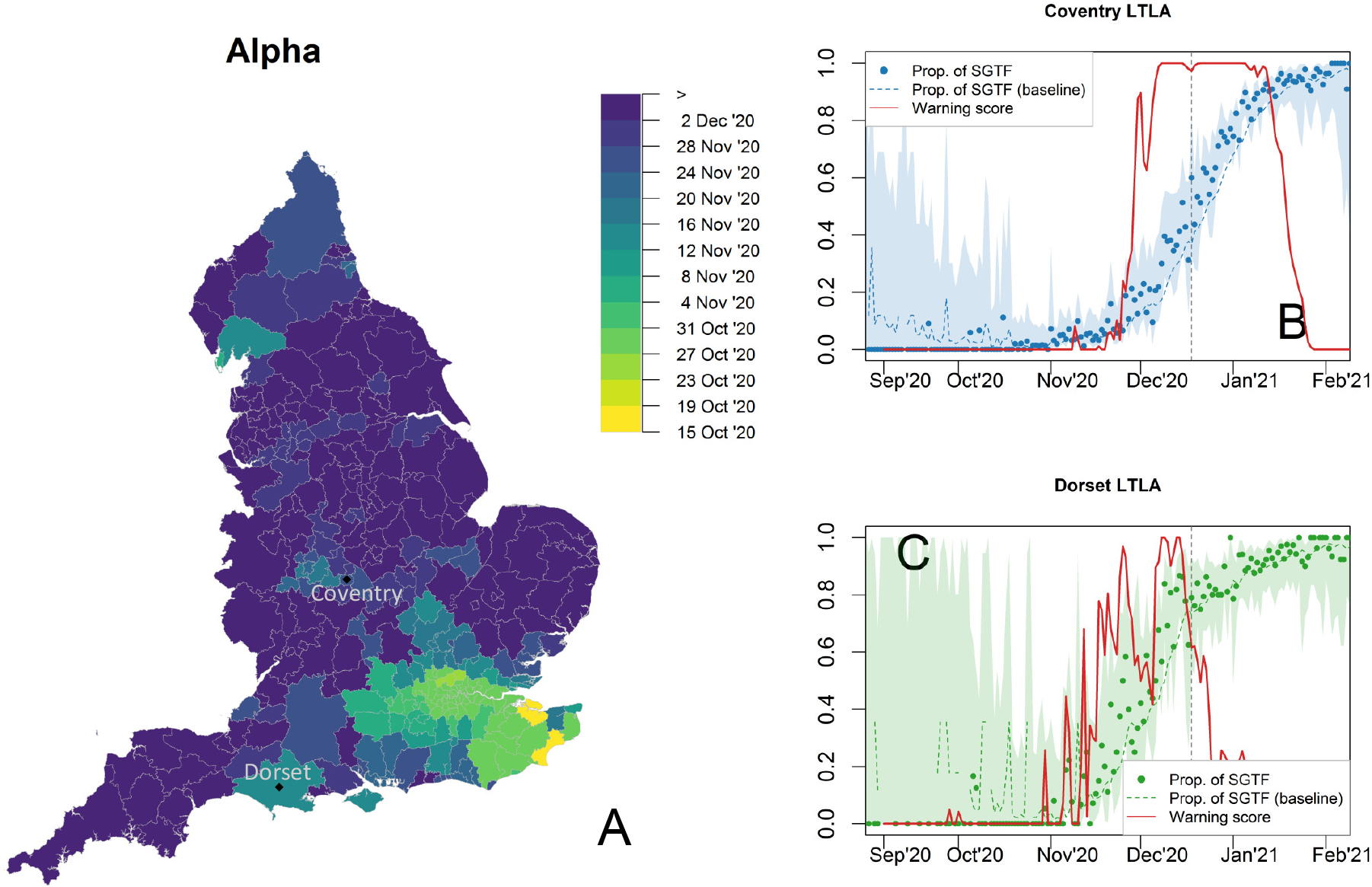
Progression of the Alpha variant and warning scores obtained with the down-sampled data set (panel analogous to figure 4 obtained with all available data). A) Spatial diffusion of warning flags for SGTFs during the Alpha invasion. The colour map from yellow to blue indicates the date when an LTLA warning score first exceeded a threshold value of 90%, starting from 15 October 2020. LTLAs in the South East are flagged first. Temporal trends in Coventry (B) and Dorset LTLAs (C). The observed proportions of SGTFs are marked by dots, with dashed lines representing null-model expectations and shaded areas 0.25%-97.5% Wilson CIs, which are wider than those in figure 4, main text. Warning scores, represented by solid red lines, only have a slightly weaker signal than those of figure 4 and in certain locations (e.g., Dorset LTLA) *w* exceeded 90% with substantial delay with respect to the full data result.

**Figure S4.**
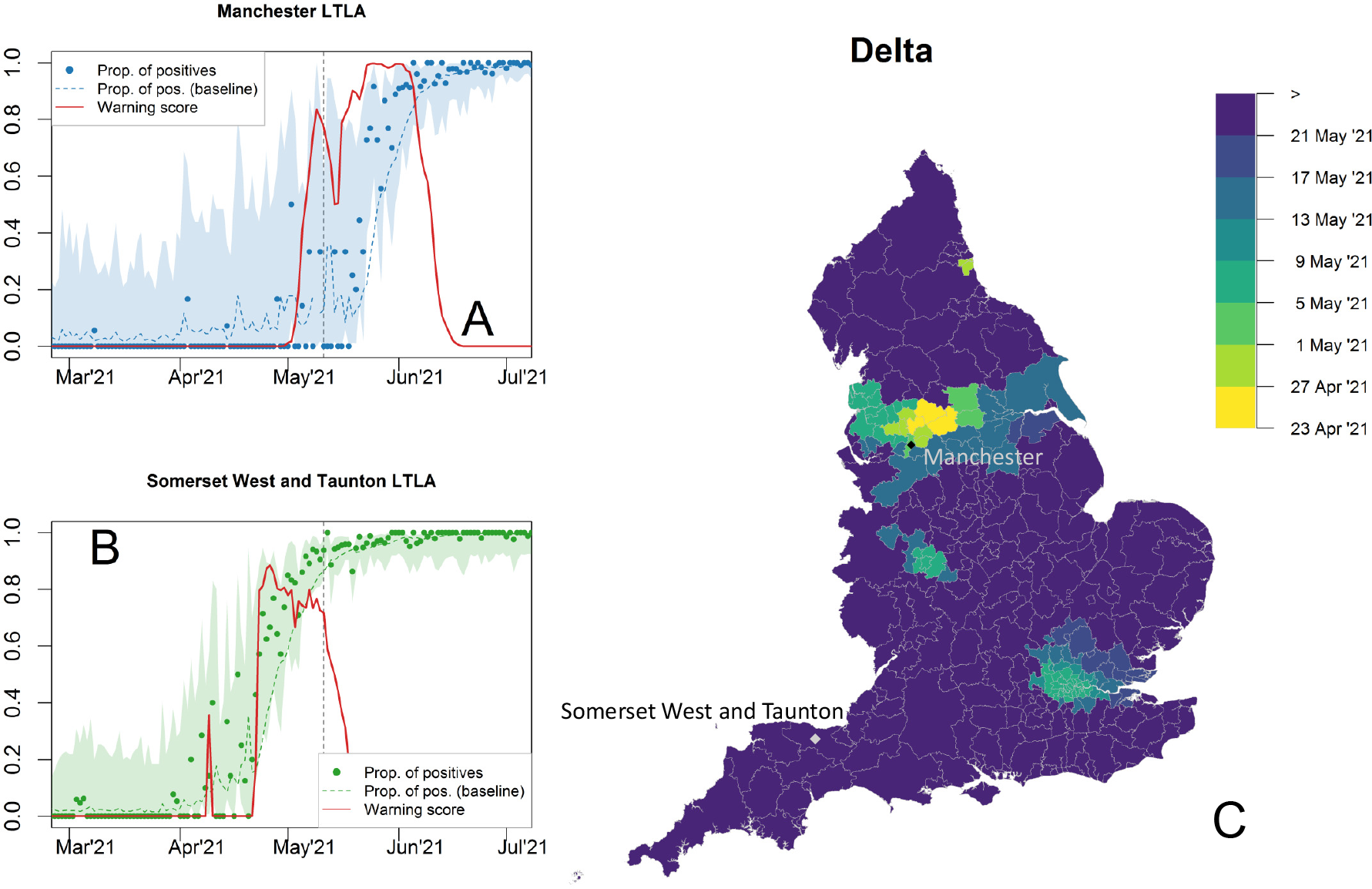
Progression of the Delta variant and warning scores for the proportion of S-gene positive tests, obtained with the down-sampled data set (panel analogous to figure 5 obtained with all available data). Temporal trends in Manchester (A) and Somerset-West-and-Taunton LTLAs (B). The observed proportions of tests are marked by dots, with dashed lines representing the expectations according to the null-model baseline. Delta invasion occurred in a phase where the total number of SARS-CoV-2 positive tests was decreasing. Low testing is exacerbated in this down-sample data and 0.25%-97.5% Wilson CIs for proportions (shaded areas) are substantially wider than those in figure 4, main text. C) Spatial diffusion of warning flags for S-gene positive tests during the Delta invasion. The colour map indicates the date when an LTLA warning score first exceeded a 90% threshold, starting from 23 April 2021. Some LTLAs never exceeded *w >* 90% (e.g., Somerset West and Taunton (B) and are coloured in dark blue.

**Figure S5.**
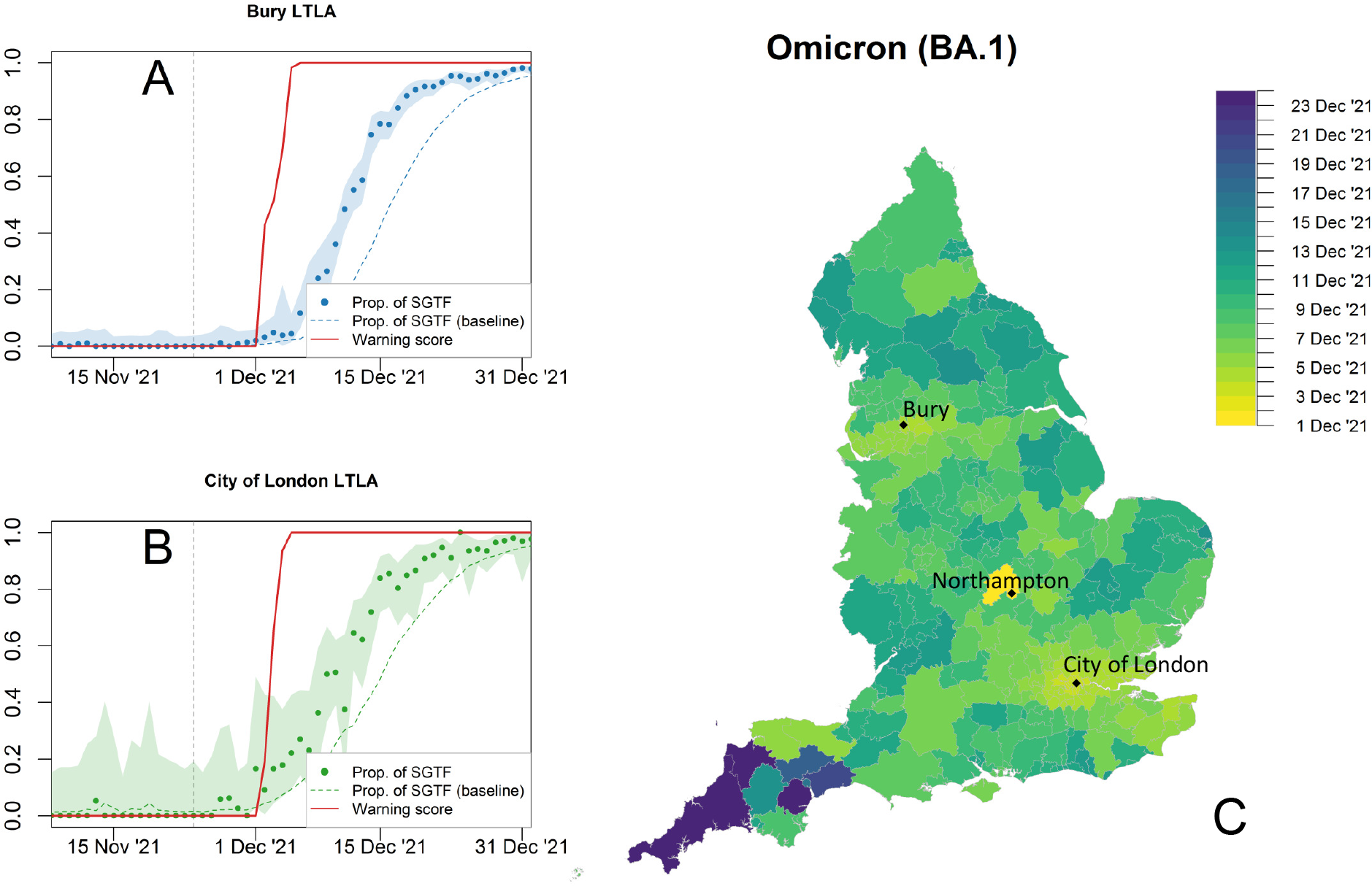
Progression of the S-gene negative Omicron variant (BA.1) and warning scores for the proportion of SGTFs, obtained with the down-sampled data set (panel analogous to figure 6, obtained with all available data, in the main text). Temporal trends and warning flag in Bury (A) and City of London (B) LTLAs.). C) Spatial diffusion of warning flags during the Omicron BA.1 invasion. The colour map indicates the date when an LTLA warning score first exceeded a threshold value of 90% starting from 1 December 2021. The areas flagged first are Manchester, Northampton, and London. Keys as in figures S3 and S3.

**Figure S6.**
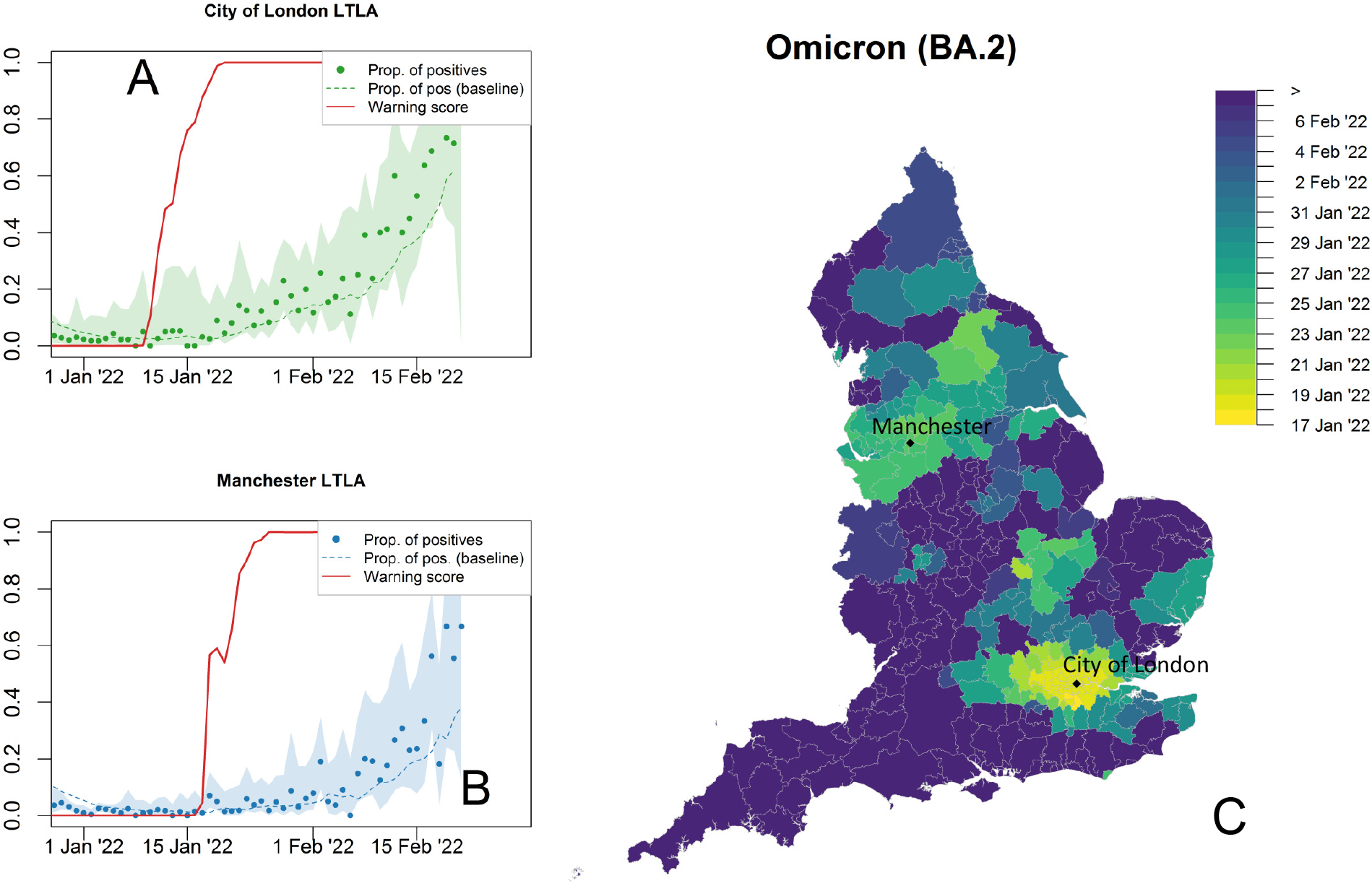
Progression of the S-gene positive Omicron variant (BA.2) and warning scores for the proportion of S-gene positive tests, obtained with the down-sampled data set (panel analogous to figure 7 obtained with all available data, in the main text). Temporal trends in City of London (A) and Manchester (B) LTLAs. C) Spatial diffusion of warning flags for SCGF during the Omicron BA.2 invasion. The colour map indicates the date when an LTLA warning score first exceeded a threshold value of 90%, starting from 17 January 2022. Keys as in figures S3 and S3.

### S4 Local anomalies of the dominant variant

Figures S7 and S8. See main text.

### S5 Warning scores from null-model data

For each lower-tier local authority (LTLA) *i* and time *t*, we simulated binomial random variables with parameters *p* and *N*_*i,t*_ (indicating the success probability and the number of trials, respectively), with *p* = 0.01, *p* = 0.1, or *p* = 0.5 (upper to low row panels in figure S9) and *N*_*i,j*_ set to the true numbers of SARS-Cov-2 tests (left panels in figure S9, “Waves”) or set to a constant value equal to the average testing capacity (right panels in figure S9, “Constant”). The application of RaNCover over these data with baseline set to *p × N*_*i,t*_ yields large warning score extremely rarely.

**Figure S7.**
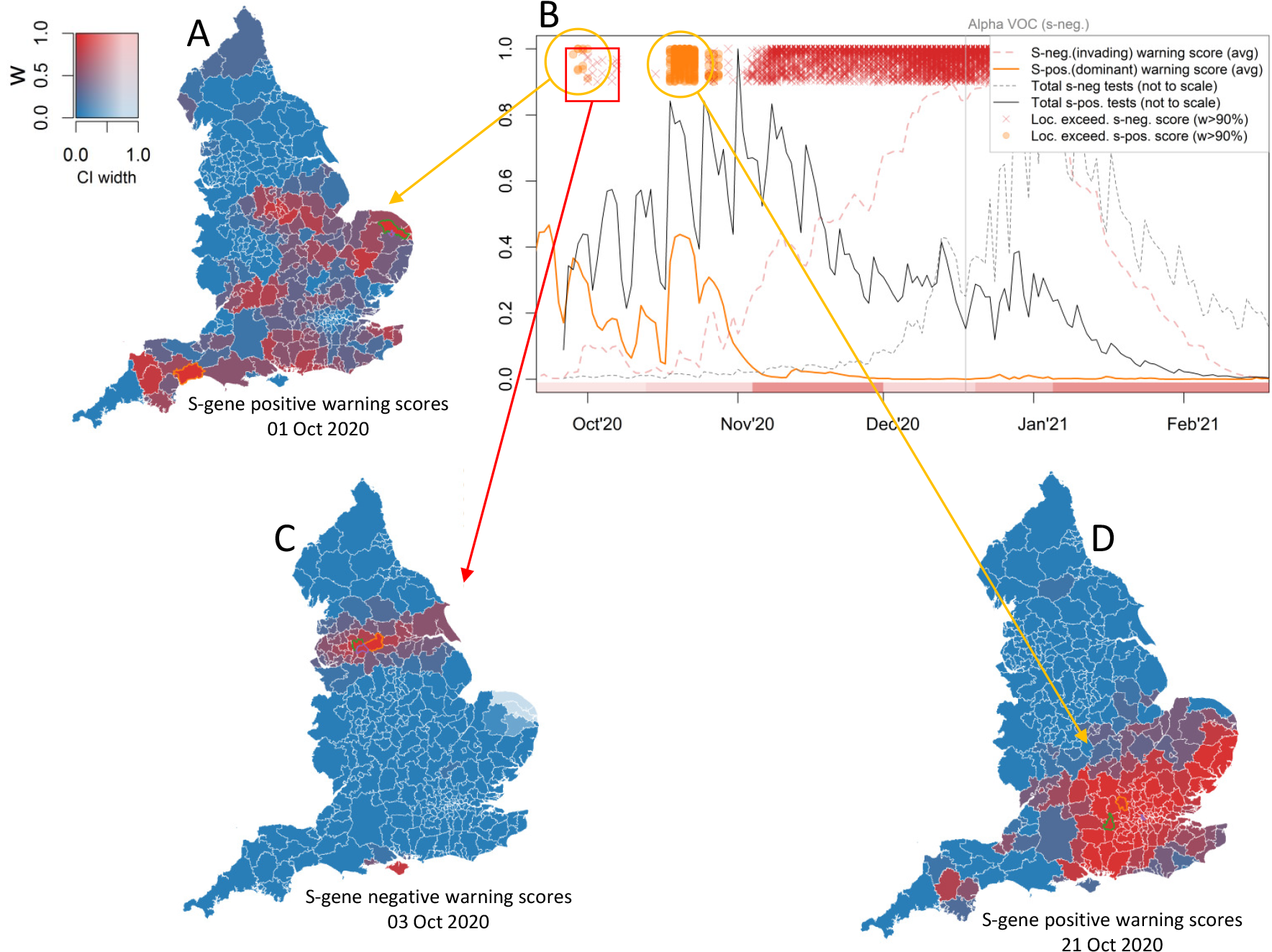
Warning flags at the beginning of the Alpha invasion. The switch from S-gene positives (here attributed to the wild-type SARS-CoV-2) to S-gene negatives (Alpha) is illustrated in panel B. The solid orange and dashed red lines are the warning scores for S-gene positives and S-gene negatives, respectively, averaged over all LTLAs. Scores for individual LTLAs that exceed the 90% threshold are illustrated as orange circles and red crosses, respectively. The total number of tests (not to scale) represented as solid and dashed grey lines, see also figure 2 in the main text. The bulk of exceeding S-gene negative warning scores corresponds to the emergence of the Alpha variant. Before that, where the wild-type is dominant, aberrant S-gene positive testing can be detected (in the regions highlighted in red, panels A and D). At the beginning of October 2020, we report a very small and short-lived cluster of S-gene negative warning scores (panel C).

**Figure S8.**
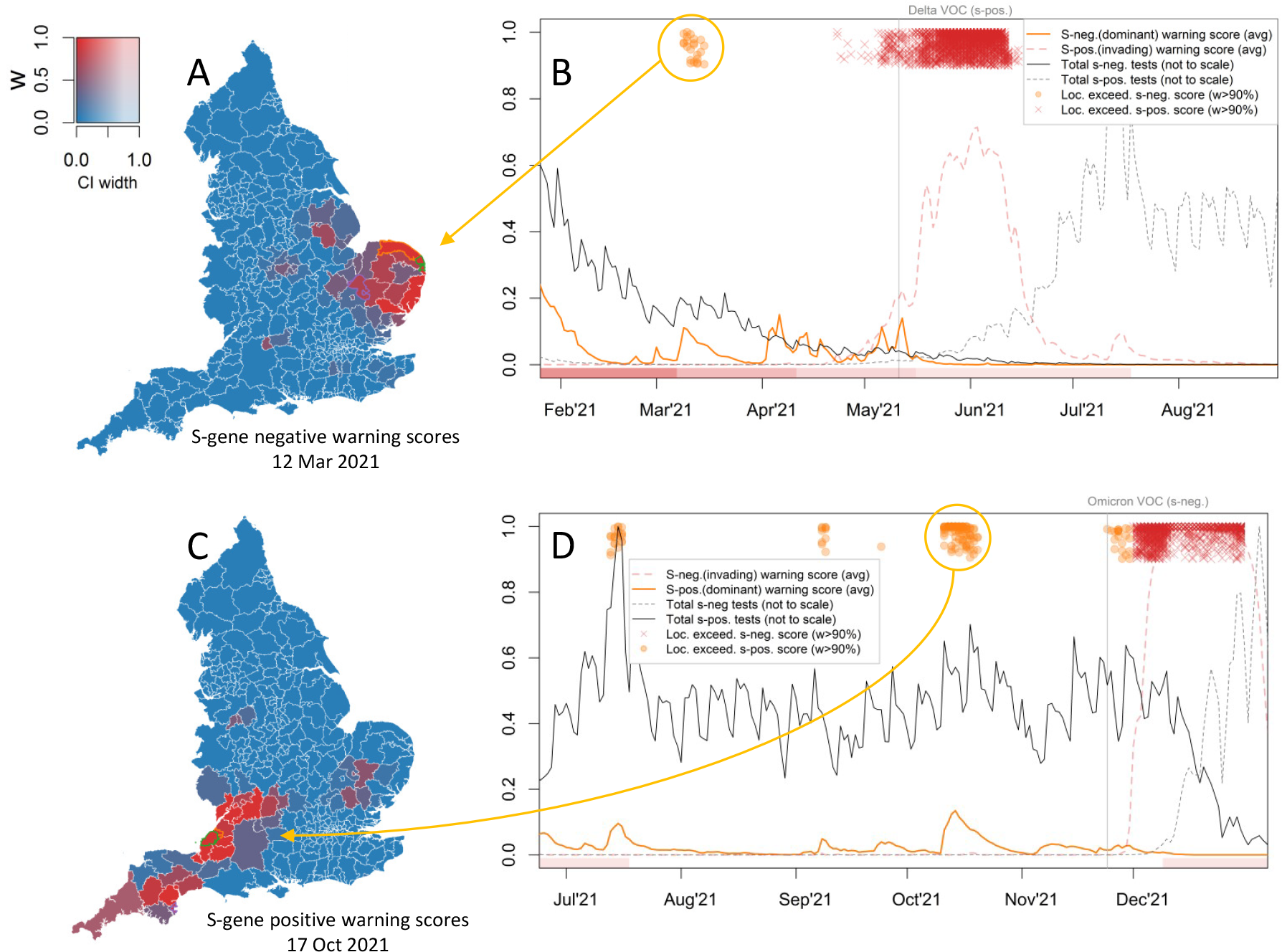
Warning flags at the beginning of the Delta and Omicron invasions. The dominant variant is Alpha in panels A and B, Delta in panels C and D. The mean warning scores (averaged over all LTLAs) are represented as solid orange and dashed red lines for the dominant and the invading variants, respectively. Locally exceeding warning scores are reported as orange circles and red crosses for the dominant and invading variants, respectively. We report exceeding warning score (*w >* 90%, orange colour) for dominant Alpha in early March in the East of England (panels A and B). Locally exceeding warning scores corresponding to the dominant Delta variant were occasionally generated from July to November 2021 (panels C and D). These detected aberrations are contextual to local maxima in the total amount of tests (solid gray lines in panels B and D, not to scale).

**Figure S9.**
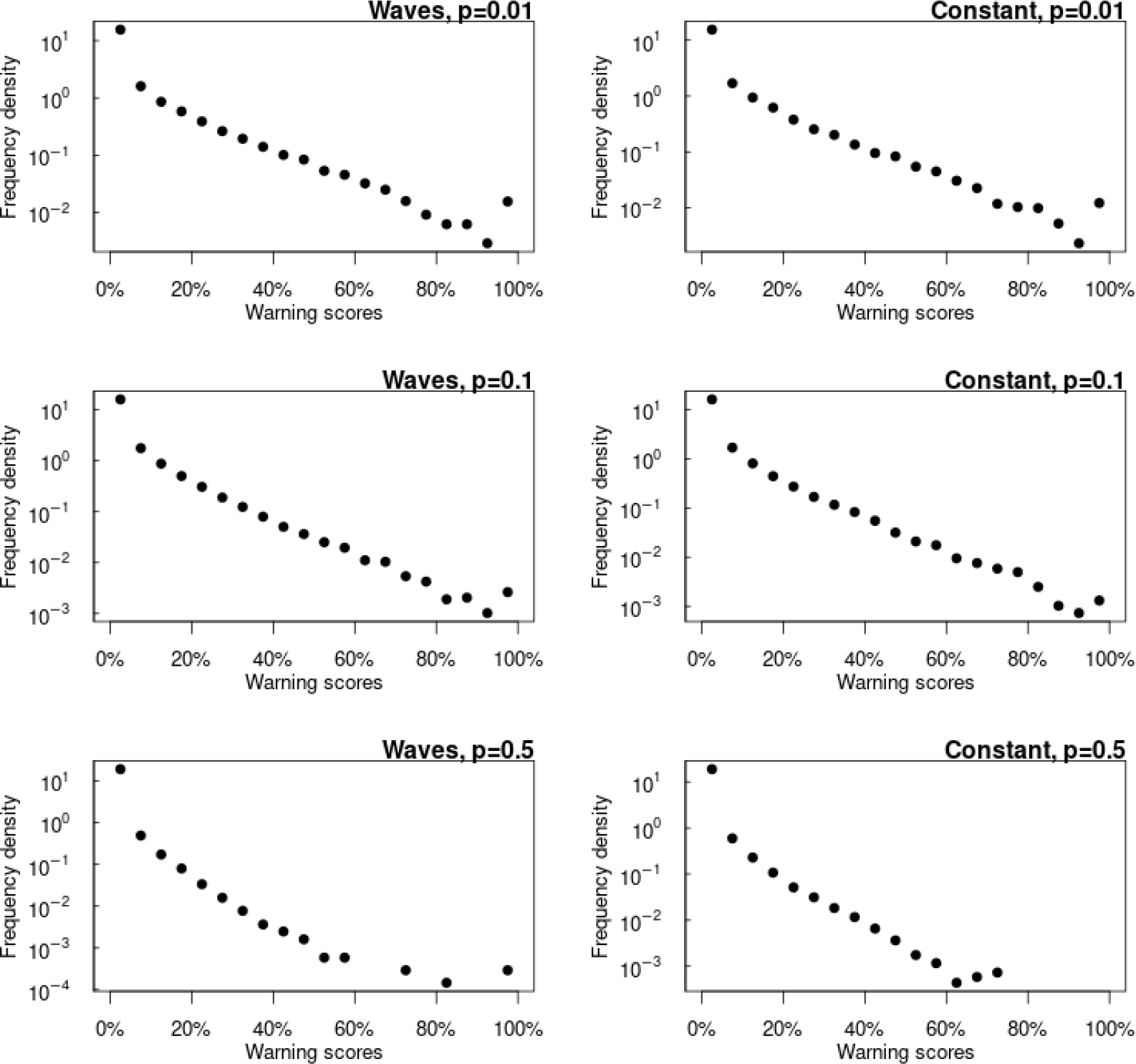
Density histograms of the warning scores obtained under six null-model assumptions.

## Notes

### Competing Interest Statement

The authors have declared no competing interest.

### Author Declarations

The ethics of the use of these data for these purposes was agreed by the UK Health Security Agency with the Government's SPI-M(O) / SAGE committees.

### Summary of Updates

Methodology and Discussion sections updated for clarity. Typos corrected. Supplementary text updated with one additional figure.

## References

1. Mathieu E, Ritchie H, Rodés-Guirao L, Appel C, Giattino C, Hasell J, et al. Coronavirus Pandemic (COVID-19). Our World in Data. 2020;.

2. World Health Organization. Public health surveillance for COVID-19: interim guidance; 2022. Available from: https://www.who.int/publications/i/item/WHO-2019-nCoV-SurveillanceGuidance-2022.2.

3. Brookmeyer R, Stroup DF. Monitoring the Health of Populations: Statistical Principles and Methods for Public Health Surveillance. New York, NY: Oxford University Press; 2004.

4. Davies NG, Kucharski AJ, Eggo RM, Gimma A, Edmunds WJ, Jombart T, et al. Effects of non-pharmaceutical interventions on COVID-19 cases, deaths, and demand for hospital services in the UK: a modelling study. The Lancet Public Health. 2020;5(7):e375–e385. doi:10.1016/S2468-2667(20)30133-X.

5. Thompson RN. Epidemiological models are important tools for guiding COVID-19 interventions. BMC Medicine. 2020;18(1):152. doi:10.1186/s12916-020-01628-4.

6. Robishaw JD, Alter SM, Solano JJ, Shih RD, DeMets DL, Maki DG, et al. Genomic surveillance to combat COVID-19: challenges and opportunities. The Lancet Microbe. 2021;2(9):e481–e484. doi:10.1016/S2666-5247(21)00121-X.

7. World Health Organization. Tracking SARS-CoV-2 variants; 2021. Available from: https://www.who.int/activities/tracking-SARS-CoV-2-variants.

8. Robertson C, Nelson TA, MacNab YC, Lawson AB. Review of methods for space–time disease surveillance. Spatial and Spatio-temporal Epidemiology. 2010;1(2-3):105–116. doi:10.1016/j.sste.2009.12.001.

9. Unkel S, Farrington CP, Garthwaite PH, Robertson C, Andrews N. Statistical methods for the prospective detection of infectious disease outbreaks: A review. Journal of the Royal Statistical Society Series A: Statistics in Society. 2012;175(1):49–82. doi:10.1111/j.1467-985X.2011.00714.x.

10. 1. Enki DG, Garthwaite PH, Farrington CP, Noufaily A, Andrews NJ, Charlett A. Comparison of Statistical Algorithms for the Detection of Infectious Disease Outbreaks in Large Multiple Surveillance Systems. PLOS ONE. 2016;11(8):e0160759. doi:10.1371/journal.pone.0160759.

11. Noufaily A, Morbey RA, Colón-González FJ, Elliot AJ, Smith GE, Lake IR, et al. Comparison of statistical algorithms for daily syndromic surveillance aberration detection. Bioinformatics. 2019;(January):1–9. doi:10.1093/bioinformatics/bty997.

12. Drake JM, Brett TS, Chen S, Epureanu BI, Ferrari MJ, Marty E, et al. The statistics of epidemic transitions. PLoS Computational Biology. 2019;15(5). doi:10.1371/journal.pcbi.1006917.

13. Brett T, Ajelli M, Liu QH, Krauland MG, Grefenstette JJ, Van Panhuis WG, et al. Detecting critical slowing down in high-dimensional epidemiological systems. PLOS Computational Biology. 2020;16(3):e1007679. doi:10.1371/JOURNAL.PCBI.1007679.

14. Southall E, Brett TS, Tildesley MJ, Dyson L. Early warning signals of infectious disease transitions: A review. Journal of the Royal Society Interface. 2021;18(182). doi:10.1098/rsif.2021.0555.

15. Southall E, Tildesley MJ, Dyson L. How early can an upcoming critical transition be detected? medRxiv. 2022; p. 2022.05.27.22275693. doi:10.1101/2022.05.27.22275693.

16. Cavallaro M, Coelho J, Ready D, Decraene V, Lamagni T, McCarthy ND, et al. Cluster detection with random neighbourhood covering: Application to invasive Group A Streptococcal disease. PLOS Computational Biology. 2022;18(11):e1010726. doi:10.1371/journal.pcbi.1010726.

17. Farrington CP, Andrews NJ, Beale AD, Catchpole MA. A Statistical Algorithm for the Early Detection of Outbreaks of Infectious Disease. Journal of the Royal Statistical Society Series A (Statistics in Society). 1996;159(3):547. doi:10.2307/2983331.

18. Yoneoka D, Kawashima T, Makiyama K, Tanoue Y, Nomura S, Eguchi A. Geographically weighted generalized Farrington algorithm for rapid outbreak detection over short data accumulation periods. Statistics in Medicine. 2021;40(28):6277–6294. doi:10.1002/sim.9182.

19. Hastie T, Tibshirani R, Friedman J. The Elements of Statistical Learning. Springer Series in Statistics. New York, NY: Springer New York; 2009. Available from: http://link.springer.com/10.1007/978-0-387-84858-7.

20. Brown KA, Gubbay J, Hopkins J, Patel S, Buchan SA, Daneman N, et al. S-Gene Target Failure as a Marker of Variant B.1.1.7 Among SARS-CoV-2 Isolates in the Greater Toronto Area, December 2020 to March 2021. JAMA. 2021;325(20):2115–2116. doi:10.1001/JAMA.2021.5607.

21. Challen R, Dyson L, Overton CE, Guzman-Rincon LM, Hill EM, Stage HB, et al. Early epidemiological signatures of novel SARS-CoV-2 variants: establishment of B.1.617.2 in England. medRxiv. 2021; p. 2021.06.05.21258365. doi:10.1101/2021.06.05.21258365.

22. Walker AS, Vihta KD, Gethings O, Pritchard E, Jones J, House T, et al. Tracking the Emergence of SARS-CoV-2 Alpha Variant in the United Kingdom. New England Journal of Medicine. 2021;385(27):2582–2585. doi:10.1056/NEJMc2103227.

23. Davies NG, Abbott S, Barnard RC, Jarvis CI, Kucharski AJ, Munday JD, et al. Estimated transmissibility and impact of SARS-CoV-2 lineage B.1.1.7 in England. Science. 2021;372(6538). doi:10.1126/science.abg3055.

24. Mishra S, Mindermann S, Sharma M, Whittaker C, Mellan TA, Wilton T, et al. Changing composition of SARS-CoV-2 lineages and rise of Delta variant in England. EClinicalMedicine. 2021;39:101064. doi:10.1016/j.eclinm.2021.101064.

25. Keeling MJ, Dyson L, Tildesley MJ, Hill EM, Moore S. Comparison of the 2021 COVID-19 roadmap projections against public health data in England. Nature Communications. 2022;13(1):4924. doi:10.1038/s41467-022-31991-0.

26. Public Health England. Investigation of novel SARS-COV-2 variant Variant of Concern 202012/01 - Technical briefing 1; 2020. Available from: https://assets.publishing.service.gov.uk/government/uploads/system/uploads/attachment_data/file/959438/Technical_Briefing_VOC_SH_NJL2_SH2.pdf.

27. Public Health England. SARS-CoV-2 variants of concern and variants under investigation in England - Technical briefing 10; 2021. Available from: https://assets.publishing.service.gov.uk/government/uploads/system/uploads/attachment_data/file/984274/Variants_of_Concern_VOC_Technical_Briefing_10_England.pdf.

28. UK Health Security Agency. SARS-CoV-2 variants of concern and variants under investigation in England - Technical briefing 30; 2021. Available from: https://assets.publishing.service.gov.uk/government/uploads/system/uploads/attachment_data/file/1038404/Technical_Briefing_30.pdf.

29. Wallinga J, Lipsitch M. How generation intervals shape the relationship between growth rates and reproductive numbers. Proceedings of the Royal Society B: Biological Sciences. 2007;274(1609):599–604. doi:10.1098/rspb.2006.3754.

30. Lehtinen S, Ashcroft P, Bonhoeffer S. On the relationship between serial interval, infectiousness profile and generation time. Journal of The Royal Society Interface. 2021;18(174):20200756. doi:10.1098/rsif.2020.0756.

31. Hart WS, Abbott S, Endo A, Hellewell J, Miller E, Andrews N, et al. Inference of the SARS-CoV-2 generation time using UK household data. eLife. 2022;11. doi:10.7554/eLife.70767.

32. Timeline of UK government coronavirus lockdowns and restrictions; 2023. Available from: https://www.instituteforgovernment.org.uk/charts/uk-government-coronavirus-lockdowns.

33. Rambaut A, Holmes EC, O’Toole A, Hill V, McCrone JT, Ruis C, et al. Addendum: A dynamic nomenclature proposal for SARS-CoV-2 lineages to assist genomic epidemiology. Nature Microbiology. 2021;6(3):415–415. doi:10.1038/s41564-021-00872-5.

34. Rambaut A, Loman N, Pybus OG, Barclay W, Barrett JC, Carabelli A, et al. Preliminary genomic characterisation of an emergent SARS-CoV-2 lineage in the UK defined by a novel set of spike mutations. 2020. Available from: https://virological.org/t/preliminary-genomic-characterisation-of-an-emergent-sars-%0Acov-2-lineage-in-the-uk-defined-by-a-novel-set-of-spike-mutations/563.

35. The COVID-19 Genomics UK (COG-UK) consortium. An integrated national scale SARS-CoV-2 genomic surveillance network. The Lancet Microbe. 2020;1(3):e99–e100. doi:10.1016/S2666-5247(20)30054-9.

36. Kraemer MUG, Hill V, Ruis C, Dellicour S, Bajaj S, McCrone JT, et al. Spatiotemporal invasion dynamics of SARS-CoV-2 lineage B.1.1.7 emergence. Science. 2021;373(6557):889–895. doi:10.1126/science.abj0113.

37. McCrone JT, Hill V, Bajaj S, Pena RE, Lambert BC, Inward R, et al. Context-specific emergence and growth of the SARS-CoV-2 Delta variant. Nature. 2022;610(7930):154–160. doi:10.1038/s41586-022-05200-3.

38. UK Cabinet Office. COVID-19 Response—Spring 2021 (Summary); 2021. Available from: https://www.gov.uk/government/publications/covid-19-response-spring-2021/covid-19-response-spring-2021-summary.

39. Ferguson NM. B.1.617.2 transmission in England: risk factors and transmission advantage; 2021. Available from: https://www.gov.uk/government/publications/imperial-college-london-delta-b16172-transmission-in-england-risk-factors

40. Keeling MJ, Brooks-Pollock E, Challen RJ, Danon L, Dyson L, Gog JR, et al. Short-term Projections based on Early Omicron Variant Dynamics in England. medRxiv. 2021;(November):2021.12.30.21268307.

41. Mahase E. Covid-19: What do we know about omicron sublineages? BMJ. 2022;376:o358. doi:10.1136/bmj.o358.

42. Statement on Omicron sublineage BA.2;. Available from: https://www.who.int/news/item/22-02-2022-statement-on-omicron-sublineage-ba.2.

43. UK Health Security Agency. COVID-19 variants identified in the UK – latest updates - GOV.UK; 2023. Available from: https://www.gov.uk/government/news/covid-19-variants-identified-in-the-uk-latest-updates.

44. Harvey WT, Carabelli AM, Jackson B, Gupta RK, Thomson EC, Harrison EM, et al. SARS-CoV-2 variants, spike mutations and immune escape. Nature Reviews Microbiology. 2021;19(7):409–424. doi:10.1038/s41579-021-00573-0.

45. UK Health Security Agency. Changes to COVID-19 testing in England from 1 April; 2022. Available from: https://www.gov.uk/government/news/changes-to-covid-19-testing-in-england-from-1-april.

46. Lo SW, Jamrozy D. Genomics and epidemiological surveillance. Nature Reviews Microbiology. 2020;18(9):478–478. doi:10.1038/s41579-020-0421-0.

47. Emma B Hodcroft. CoVariants: SARS-CoV-2 Mutations and Variants of Interest. 2021. Available from: https://covariants.org/.

48. Centers for Disease Control and Prevention. What is Genomic Surveillance?; 2022. Available from: https://www.cdc.gov/coronavirus/2019-ncov/variants/genomic-surveillance.html.

49. Wellcome Sanger Institute’s COVID-19 Genomics Initiative. COVID-19 Genomic Surveillance; 2023. Available from: https://covid19.sanger.ac.uk.

50. Keeling MJ, Rohani P. Modeling infectious diseases in humans and animals. Princeton, New Jersey: Princeton University Press; 2008.

51. Iacobucci G. Covid-19: Three tier alert system takes effect across England. BMJ. 2020;371:m3961. doi:10.1136/bmj.m3961.

52. Buckeridge DL, Burkom H, Campbell M, Hogan WR, Moore AW. Algorithms for rapid outbreak detection: a research synthesis. Journal of Biomedical Informatics. 2005;38(2):99–113. doi:10.1016/J.JBI.2004.11.007.

53. Pebesma E. Simple Features for R: Standardized Support for Spatial Vector Data. The R Journal. 2018;10(1):439. doi:10.32614/RJ-2018-009.

